# Recruitment Success Among Ethnic Minority Doctors and International Medical Graduates in UK Medical Specialty Recruitment: A Repeated Cross-Sectional Analysis of NHS England Data (2021–2025)

**DOI:** 10.64898/2026.10.06.26364840

**Authors:** Ryan Malcolm Hum, Hannah Aaria Shafi, Seema D Sharma, Lysette Michele Marshall

## Abstract

**Background:** Ethnic minority doctors and international medical graduates (IMGs) now make up most applicants to UK specialty training. Earlier work has reported lower recruitment success for some ethnic groups, but evidence that covers recent cycles, every training level, all recruitment stages and disaggregated ethnic groups is limited. We describe trends in ethnicity and country of primary medical qualification among applicants to UK specialty training between 2021 and 2025, and compare the odds of receiving a training offer between groups.

**Methods:** Repeated cross-sectional analysis of national specialty recruitment data published by NHS England for 2021–2025, comprising 328,839 applications to entry-level (ST1/CT1) and higher specialty (ST3+) programmes. Unadjusted odds ratios (ORs) with 95% confidence intervals (CIs) for receiving an offer were calculated for each programme with at least 500 cumulative offers, comparing broad ethnic groups with White applicants, sub-ethnic groups within broad groups, and EU/EEA and rest-of-world graduates with UK graduates. Spearman’s rank correlation tested whether programmes with more rest-of-world applicants showed larger ethnic differences. Ethnicity offers for 2023 were excluded because the published 2023 tables duplicate those for 2021.

**Results:** Applications rose 3.1-fold, from 36,709 in 2021 to 114,557 in 2025, while annual offers stayed between 16,988 and 19,710. The share of applications from White applicants fell from 34.9% to 16.0%, and the share from doctors qualifying outside the UK and EU rose from 43.9% to 69.4%. Asian and Black applicants had lower offer odds than White applicants in every programme analysed (9 entry-level and 12 higher programmes), with the largest differences at entry level in Anaesthetics (Black vs White OR 0.027, 95% CI 0.020–0.036) and Emergency Medicine (Asian vs White OR 0.086, 95% CI 0.075–0.098). Chinese applicants had lower odds in 4 of 9 entry-level and 1 of 12 higher programmes. Within broad groups, Pakistani and Bangladeshi applicants had lower odds than Indian applicants, and Black Caribbean applicants higher odds than Black African applicants, at both levels. Rest-of-world graduates had lower odds than UK graduates in all 11 entry-level and 17 higher programmes. A programme’s share of rest-of-world applicants was not significantly correlated with Asian or Black offer ORs (*ρ −*0.45 to *−*0.27; all *p ≥* 0.22).

**Conclusions:** Centralised medical recruitment in the UK exhibits some persistent ethnic and international qualification disparities despite rising application diversity. The findings highlight the necessity of disaggregating ethnic groups and addressing subjective evaluation vulnerabilities within multi-stage recruitment processes to foster equity.

## Background

The United Kingdom is ethnically diverse. In the 2021 Census of England and Wales, 81.7% of the population identified as White, 9.3% as Asian, 4.0% as Black, 2.9% as Mixed and 0.7% as Chinese [1]. The NHS is the largest employer in the UK, employing over 1.3 million people, of whom 25.7% are from non-White ethnic groups [2]. Of the 125,568 doctors employed in the NHS, 50% were from White ethnic backgrounds, 32% from Asian, 6% from Black and 3% from Chinese ethnic backgrounds [2, 3].

A workforce that reflects the population it serves matters for patients and for the health service. Diverse teams have been associated with better problem solving, communication and patient outcomes [4], and doctors who can progress according to their ability are more likely to remain in the workforce. Yet differences in career progression by ethnicity are well documented. Doctors from ethnic minority backgrounds have lower attainment in undergraduate and postgraduate assessments [5], describe barriers in training relationships and assessment [6], and are under-represented in senior roles across the NHS [7]. Over 30 years ago, applications for senior house officer posts with Asian names were shown to be less likely to be shortlisted than otherwise identical applications with English names [8].

Entry to specialty training in the UK is through national recruitment processes that are intended to be fair and transparent. The number of UK medical school places is capped by government and managed by national bodies such as the Office for Students and the Department of Health and Social Care [9, 10], in line with workforce policy such as the NHS Long Term Workforce Plan [11]. In 2019 the Resident Labour Market Test for medical specialty training was lifted when all medical practitioners were added to the Shortage Occupation List, so that international medical graduates (IMGs) could apply for UK specialty training on equal terms with UK graduates [12]. Applications from IMGs then rose sharply, competition ratios for many specialties increased [13, 14], and IMGs now form a large part of the medical workforce [15]. In 2026 the Medical Training (Prioritisation) Act was passed, which again prioritises UK graduates for specialty training places [16]. The Act treats country of qualification, rather than ethnicity, as the policy lever; this is important for interpretation, because IMGs are more likely than UK graduates to be from ethnic minority groups, and country of qualification is itself associated with progression through training and success in specialty recruitment [17, 18].

Previous studies have reported that doctors from some ethnic groups, and IMGs, are less likely to be offered training posts in certain specialties [3, 6, 17–20]. Aggarwal and colleagues used freedom of information requests to describe the diversity of successful applicants in 2021–2022 and protected-characteristic outcomes in 2021–2024, including country of graduation [17, 18]; Brown and colleagues described ethnic differences in specialty destinations using a repeated cross-sectional design [20]. We extend this work using the national recruitment data that NHS England publishes for every specialty and level, adding the 2025 recruitment round, all three recruitment stages (application, offer and acceptance), disaggregated sub-ethnic groups, and country of qualification.

Our aims were to describe trends in self-reported ethnicity and country of qualification among applicants to UK specialty training between 2021 and 2025, to describe how representation varies between specialties and recruitment stages, and to compare the odds of receiving an offer between ethnic groups, between sub-ethnic groups, and between UK graduates and IMGs.

## Methods

### Study design and data source

We conducted a repeated cross-sectional analysis of national specialty training recruitment data for the 2021 to 2025 recruitment years. NHS England publishes, for each recruitment year, tables of applications made, appointable applicants, offers made and offers accepted, broken down by programme and entry level, for self-reported ethnicity and for country of primary medical qualification [21]. These programmes are recruited nationally for posts across the UK. Ethnicity and country of qualification are reported in separate tables and cannot be linked. Counts refer to applications, not individual applicants: one doctor applying to several programmes contributes one application to each.

### Data verification

Before analysis we checked the downloaded tables against the NHS England web pages (snapshots saved 10 November 2026). The 2023 ethnicity tables for offers made and offers accepted published by NHS England are cell-for-cell identical to the 2021 tables, including the 2021 programme list and category labels, whereas the 2023 applications table is specific to 2023. We therefore excluded 2023 ethnicity offers and acceptances; 2023 ethnicity applications are reported descriptively but, because offer odds are calculated only from programme-years with both applications and offers, 2023 does not contribute to any ethnicity odds ratio.

### Programmes and training levels

Programme titles were harmonised across years to account for changes in nomenclature. Acute Care Common Stem routes were combined with their parent specialty (for example, ACCS Emergency Medicine with Emergency Medicine, and ACCS Internal Medicine with Internal Medicine Training). Programmes were grouped by entry level into entry-level training (ST1/CT1, including CT1 and ST1/2 entry points) and higher specialty training (ST3 and above). In total, 61 programmes were represented.

### Ethnicity and country of qualification

Ethnicity was self-reported using 16 categories plus ‘not stated’. Following the categories in the published data, these were grouped into six broad groups: White (British, Irish, any other White background), Asian (Indian, Pakistani, Bangladeshi, any other Asian background), Black (African, Caribbean, any other Black background), Mixed (White and Asian, White and Black African, White and Black Caribbean, any other mixed background), Chinese, and any other ethnic group. Chinese applicants are reported as a separate category in the source data and were analysed separately. Because broad groups can hide differences between their members [22, 23], we also compared sub-ethnic groups within the White, Asian and Black groups, using the largest subgroup as the reference (White British, Indian and Black African, respectively). Applications with ethnicity not stated were described but excluded from comparisons. Country of primary medical qualification was reported as UK, EU/EEA or rest of world; IMGs were defined as EU/EEA plus rest-of-world graduates.

### Suppressed cells and sensitivity analysis

NHS England suppresses counts below five. Suppressed cells were set to 2.5, the mid-point of the possible range. To test the effect of this choice, all analyses of offer odds were repeated with suppressed cells set to 0 and to 5.

### Statistical analysis

For each programme, applications and offers were summed across recruitment years, using only programme-years for which both applications and offers were published. The odds ratio (OR) for receiving an offer was calculated from the resulting two-by-two table as the odds of an offer in the group of interest divided by the odds of an offer in the reference group, with a 95% CI calculated on the log scale (Wald method). A continuity correction of 0.5 was added to every cell only when a cell was zero. These are unadjusted ORs: no regression model was fitted and no covariates were included, because the published tables do not cross-tabulate ethnicity with any other characteristic. White applicants were the reference group for comparisons between broad ethnic groups, and UK graduates for comparisons by country of qualification. To limit imprecision, programme-level ORs were calculated only for programmes with at least 500 cumulative offers, following our previous work [24]; all programmes were retained in national totals. Composition by programme was shown for programmes with at least 500 cumulative applications, offers or acceptances at the relevant stage.

Within-group comparisons were pooled across programmes meeting the 500-offer threshold, separately for entry-level and higher training, by summing offers and applications (crude pooled OR). Because crude pooling can be confounded by differences in where subgroups apply, we also calculated Mantel–Haenszel ORs stratified by programme.

To explore whether ethnic differences were larger in programmes with more international applicants, we calculated Spearman’s rank correlation between each programme’s percentage of applications from rest-of-world graduates (2021–2025) and its Asian vs White and Black vs White offer OR, separately for entry-level and higher programmes. This is a programme-level (ecological) analysis and does not describe individual applicants.

All tests were two-sided with *α* = 0.05. We did not adjust for multiple comparisons; results are presented as estimates with confidence intervals and should be read as descriptive. Analyses were performed in Python 3.12.5 (pandas 2.2.2, NumPy 1.26.4, SciPy 1.14.1, Matplotlib 3.9.2). The study is reported following the STROBE guidance for cross-sectional studies.

## Results

### Applications by ethnicity, 2021–2025

Between 2021 and 2025 there were 328,839 applications, 269,215 to entry-level and 59,624 to higher programmes. Annual applications rose 3.1-fold, from 36,709 in 2021 to 114,557 in 2025, whereas the annual number of offers changed little (16,988 to 19,710 in the years with valid data; Supplementary Table S1). The share of applications from White applicants fell from 34.9% (n=12,797) in 2021 to 16.0% (n=18,332) in 2025. The share from Asian applicants rose from 30.7% (n=11,278) to 40.2% (n=46,052), and from Black applicants from 11.7% (n=4,300) to 22.3% (n=25,494). The share from Chinese applicants fell from 3.2% (n=1,162) to 1.6% (n=1,818). Ethnicity was not stated on 8.0% of applications in 2021 and 6.4% in 2025 (Figure 1; sub-ethnic composition in Supplementary Figure S1).

**Figure 1.**
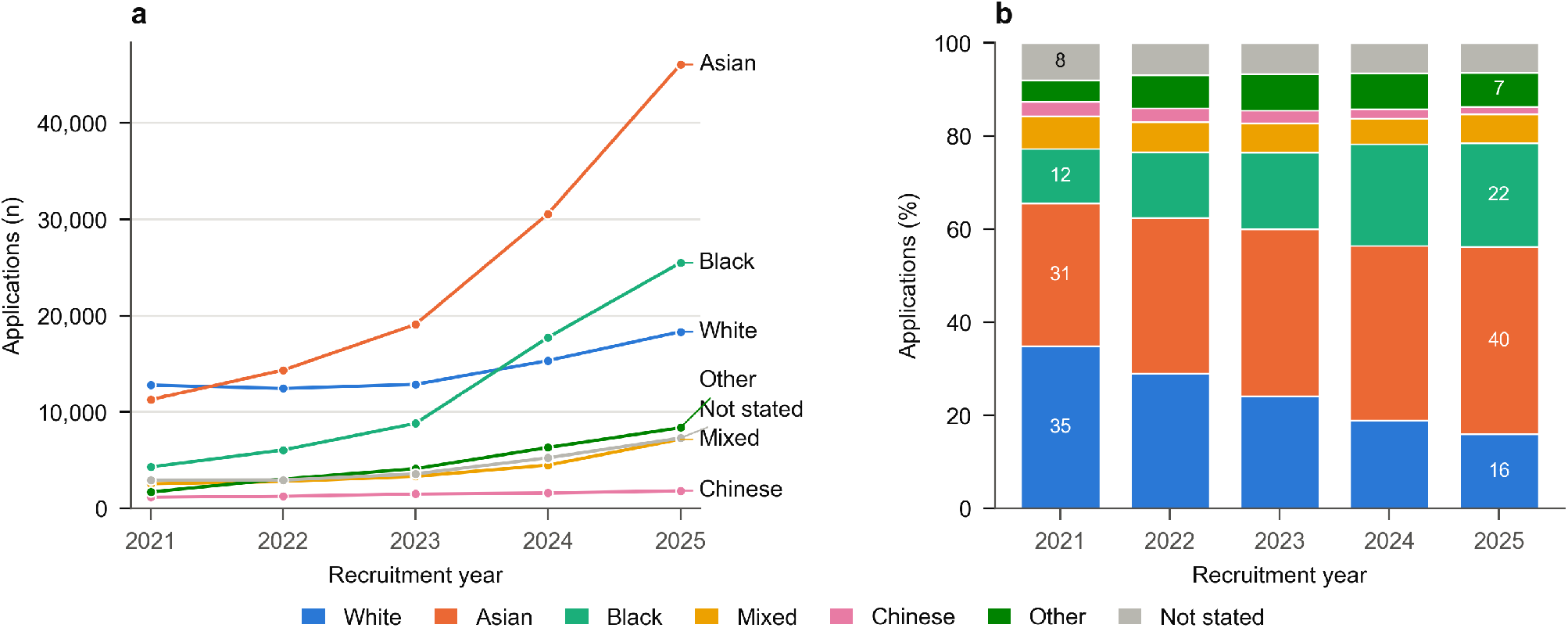
Applications to UK specialty training by self-reported ethnic group, 2021–2025. (a) Number of applications by ethnic group. (b) Percentage of applications in each ethnic group; labels show percentages for 2021 and 2025.

### Applications by country of qualification, 2021–2025

Applications from doctors who qualified outside the UK and EU rose from 16,066 in 2021 to 79,406 in 2025, and their share of applications from 43.9% to 69.4%. Applications from UK graduates also rose, from 18,200 to 31,252, but their share fell from 49.8% to 27.3%. EU/EEA graduates made up 6.3% of applications in 2021 and 3.3% in 2025. UK graduates received 65.7% of offers in 2021 and 59.0% in 2025 (Figure 2; Supplementary Table S2).

**Figure 2.**
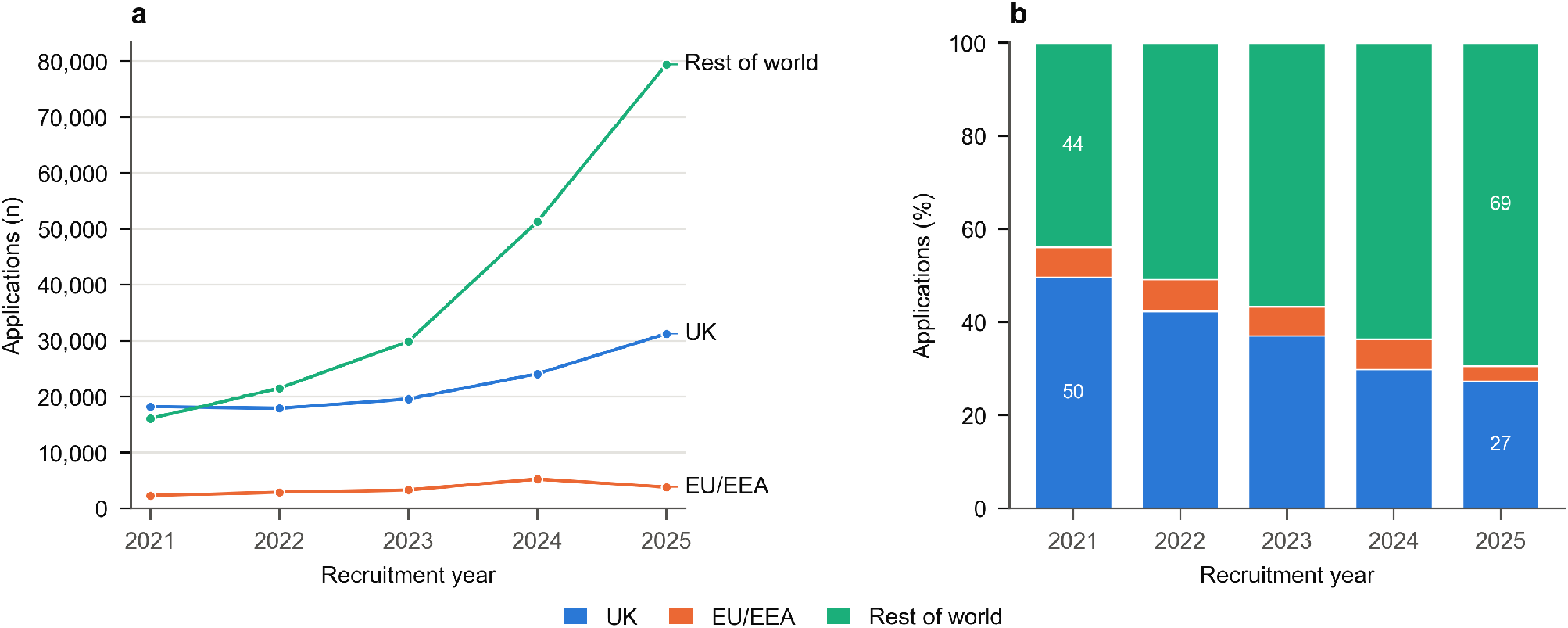
Applications to UK specialty training by country of primary medical qualification, 2021–2025. (a) Number of applications. (b) Percentage of applications; labels show percentages for 2021 and 2025.

### Ethnicity of applicants by specialty

The ethnic composition of applicants varied widely between specialties (Figure 3). At entry level, the specialties with the highest percentage of White applicants were Public Health Medicine, Anaesthetics, and Paediatrics; of Asian applicants, General Practice, Clinical Radiology, and Internal Medicine; of Black applicants, Obstetrics and Gynaecology, Community Sexual and Reproductive Health, and Core Psychiatry; and of Chinese applicants, Ophthalmology, Core Surgical Training, and Internal Medicine. At higher level, the corresponding specialties were Anaesthetics, Palliative Medicine, and Intensive Care Medicine for White applicants; Endocrinology and Diabetes Mellitus, General (Internal) Medicine, and Acute Internal Medicine for Asian applicants; Occupational Medicine, Paediatrics, and Obstetrics and Gynaecology for Black applicants; and Dermatology, Plastic Surgery, and Otolaryngology for Chinese applicants. Composition by sub-ethnic group, and at the offer and acceptance stages, is shown in Supplementary Figure S2–Supplementary Figure S6.

**Figure 3.**
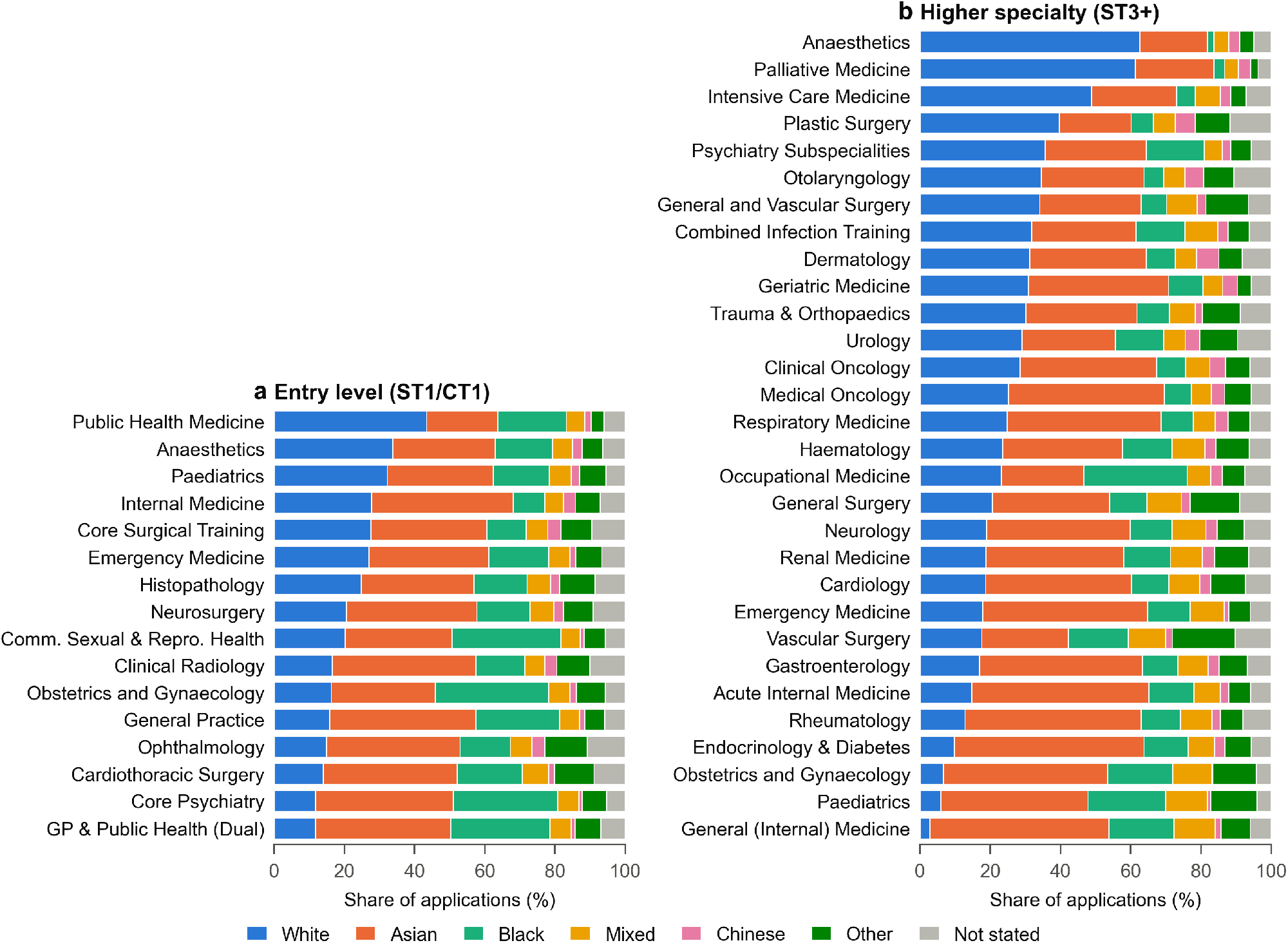
Ethnic composition of applications by specialty: (a) entry-level (ST1/CT1) and (b) higher specialty (ST3+) programmes, 2021–2025. Programmes are ordered by the percentage of White applicants. Programmes with fewer than 500 cumulative applications are not shown.

### Country of qualification of applicants by specialty

At entry level, the specialties with the highest percentage of applications from UK graduates were Public Health Medicine, Core Surgical Training, and Anaesthetics, and those with the highest percentage from IMGs were Core Psychiatry, Cardiothoracic Surgery, and General Practice and Public Health (Dual CCT). At higher level, the highest percentages of UK graduates were in Anaesthetics, Palliative Medicine, and Otolaryngology, and the highest percentages of IMGs in Obstetrics and Gynaecology, Paediatrics, and General (Internal) Medicine (Figure 4; offers and acceptances in Supplementary Figure S7 and Supplementary Figure S8).

**Figure 4.**
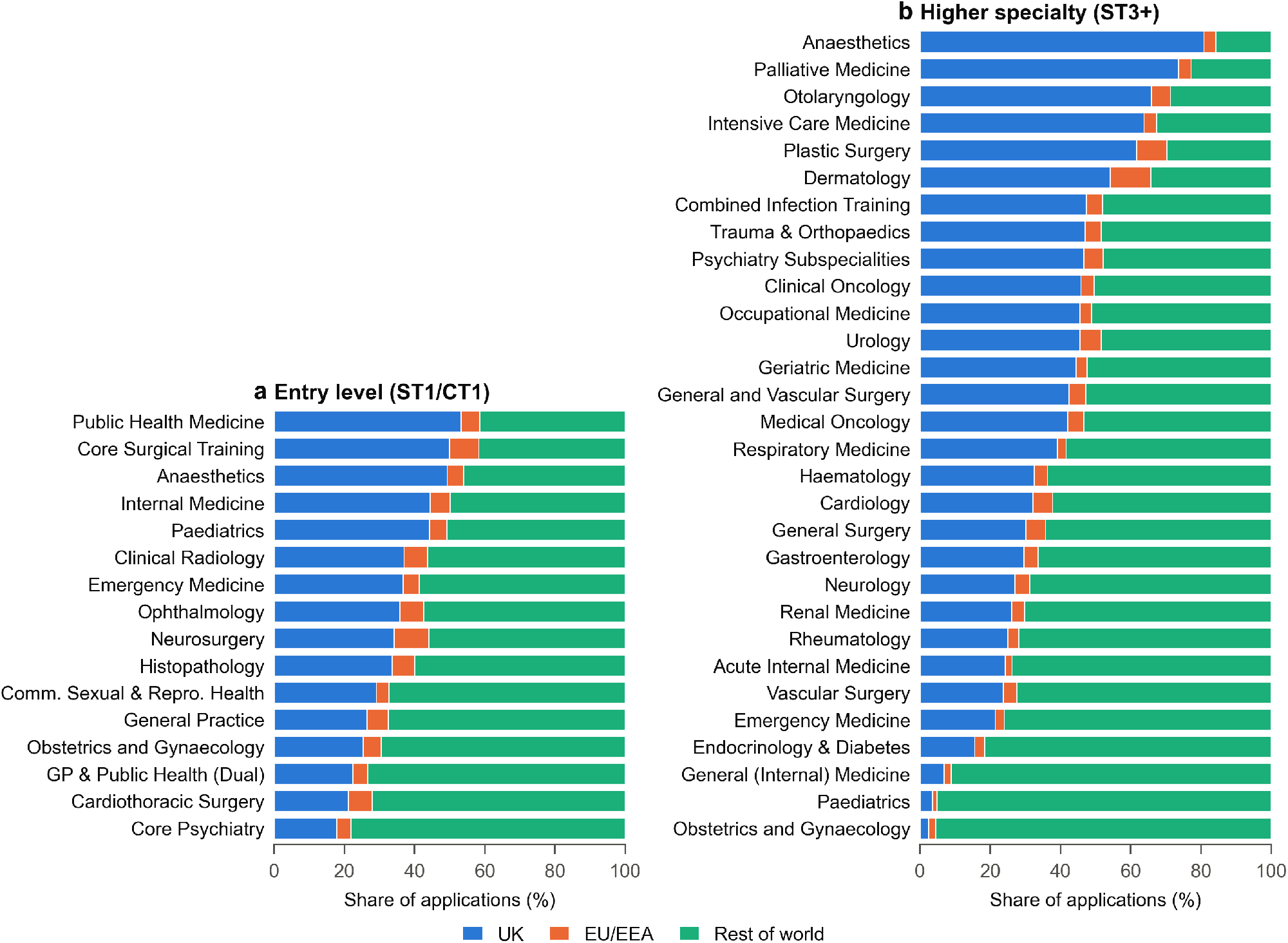
Country of primary medical qualification of applications by specialty: (a) entry-level (ST1/CT1) and (b) higher specialty (ST3+) programmes, 2021–2025. Programmes are ordered by the percentage of UK graduates. Programmes with fewer than 500 cumulative applications are not shown.

### Representation across recruitment stages

White applicants made up a larger share of offers than of applications, and Asian and Black applicants a smaller share (Supplementary Figure S13). In entry-level programmes, White applicants made up 20.1% of applications, 38.0% of offers and 38.7% of acceptances; Asian applicants 37.5%, 29.2% and 29.0%; and Black applicants 21.2%, 11.8% and 12.8%. Shares at acceptance were close to shares at offer, so differences arose between application and offer. Across all programmes, 49.1% of applications from White applicants resulted in an offer, compared with 21.5% from Asian, 15.2% from Black and 43.4% from Chinese applicants.

### Odds of receiving an offer by ethnicity

In total, 21 programmes met the 500-offer threshold (9 entry-level and 12 higher). Asian and Black applicants had significantly lower odds of receiving an offer than White applicants in every one of these programmes (Figure 5; Supplementary Table S3). At entry level, the lowest ORs for Asian applicants were in Emergency Medicine (OR 0.086, 95% CI 0.075–0.098, *p <* 0.001) and Anaesthetics (OR 0.12, 95% CI 0.11–0.14, *p <* 0.001), and for Black applicants in Anaesthetics (OR 0.027, 95% CI 0.020–0.036, *p <* 0.001) and Emergency Medicine (OR 0.053, 95% CI 0.043–0.066, *p <* 0.001). At higher level, the lowest ORs for Asian applicants were in Geriatric Medicine (OR 0.26, 95% CI 0.19–0.35, *p <* 0.001) and Trauma and Orthopaedic Surgery (OR 0.30, 95% CI 0.24–0.38, *p <* 0.001), and for Black applicants in Trauma and Orthopaedic Surgery (OR 0.17, 95% CI 0.11–0.25, *p <* 0.001) and Haematology (OR 0.20, 95% CI 0.13–0.30, *p <* 0.001).

**Figure 5.**
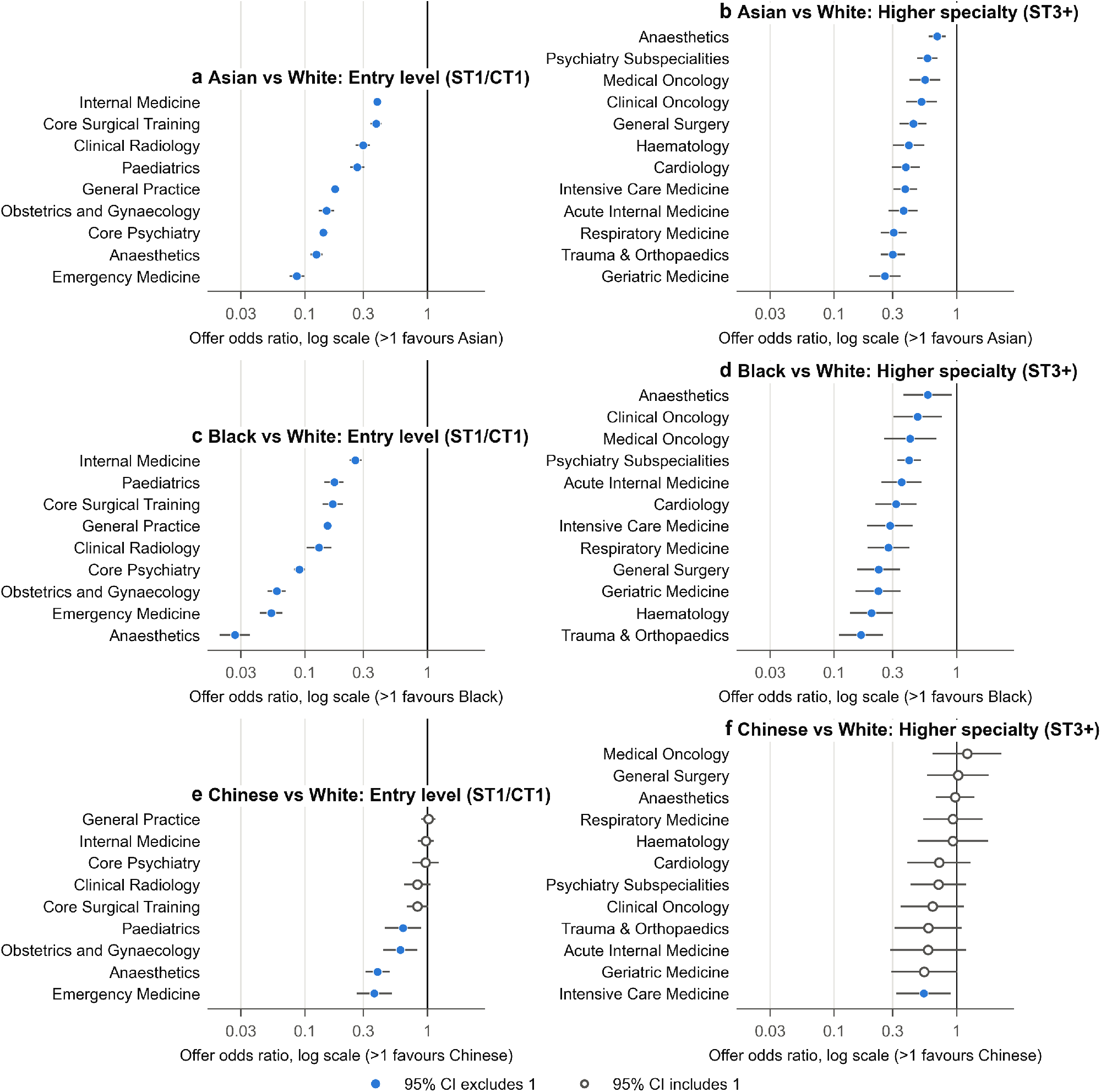
Odds of receiving a training offer for (a, b) Asian, (c, d) Black and (e, f) Chinese applicants compared with White applicants, by programme, for entry-level (left) and higher specialty (right) programmes. Points show unadjusted odds ratios and lines 95% CIs, on a log scale; values below 1 indicate lower odds of an offer than White applicants. Filled points: 95% CI excludes 1. Programmes with at least 500 cumulative offers in 2021, 2022, 2024 and 2025.

Results for Chinese applicants were more variable. Chinese applicants had significantly lower odds than White applicants in 4 of 9 entry-level programmes, most markedly in Emergency Medicine (OR 0.37, 95% CI 0.26–0.52) and Anaesthetics (OR 0.39, 95% CI 0.31–0.49), and in 1 of 12 higher programmes (Intensive Care Medicine, OR 0.54, 95% CI 0.32–0.90); in the remaining programmes the 95% CI included 1. Applicants from Mixed backgrounds had lower odds than White applicants in 9 of 9 entry-level and 11 of 12 higher programmes, and applicants from other ethnic groups in 9 of 9 and 12 of 12, respectively (Supplementary Figure S12).

### Odds of receiving an offer by sub-ethnic group

Within the Asian group, Pakistani applicants had lower odds of an offer than Indian applicants at entry level (OR 0.68, 95% CI 0.65–0.71) and higher level (OR 0.57, 95% CI 0.52–0.64), as did Bangladeshi applicants (entry level OR 0.80, 95% CI 0.74–0.86; higher level OR 0.64, 95% CI 0.52–0.78). Applicants from other Asian backgrounds had slightly higher odds than Indian applicants at entry level (OR 1.10, 95% CI 1.05–1.16) but not at higher level (OR 0.94, 95% CI 0.83–1.06). Within the Black group, Black Caribbean applicants had higher odds than Black African applicants at both levels (entry OR 2.50, 95% CI 2.15–2.90; higher OR 1.92, 95% CI 1.28–2.88); applicants from other Black backgrounds had higher odds at entry level (OR 1.75, 95% CI 1.51–2.04) but not at higher level (OR 0.77, 95% CI 0.54–1.10). Within the White group, applicants from other White backgrounds had markedly lower odds than White British applicants (entry OR 0.27, 95% CI 0.26–0.29; higher OR 0.56, 95% CI 0.49–0.62); White Irish applicants had lower odds at entry level (OR 0.75, 95% CI 0.68–0.82) but not at higher level (OR 0.89, 95% CI 0.73–1.10) (Supplementary Figure S9; Supplementary Table S5). Mantel–Haenszel estimates stratified by programme were similar in direction and significance (Supplementary Table S5).

Programme-level comparisons were possible for the three largest entry-level programmes (Supplementary Table S6). In General Practice, lower odds were seen for Bangladeshi vs Indian, Pakistani vs Indian, Irish vs British, and Other White vs British, and higher odds for Other Asian vs Indian, Caribbean vs African, and Other Black vs African. In Internal Medicine, lower odds were seen for Bangladeshi vs Indian, Pakistani vs Indian, and Other White vs British, and higher odds for Caribbean vs African. In Core Psychiatry, lower odds were seen for Bangladeshi vs Indian, Pakistani vs Indian, Irish vs British, and Other White vs British, and higher odds for Caribbean vs African.

### Odds of receiving an offer by country of qualification

Graduates from outside the UK and EU had significantly lower odds of an offer than UK graduates in all 11 entry-level programmes (ORs 0.018–0.25) and all 17 higher programmes (ORs 0.094–0.53) meeting the threshold (Supplementary Figure S10; Supplementary Table S4). The largest differences were in entry-level Anaesthetics (OR 0.018, 95% CI 0.015–0.022) and Ophthalmology (OR 0.043, 95% CI 0.031–0.059), and in higher-level Trauma and Orthopaedic Surgery (OR 0.094, 95% CI 0.076–0.116) and Emergency Medicine (OR 0.114, 95% CI 0.090–0.145). EU/EEA graduates also had lower odds than UK graduates in all 11 entry-level programmes and in 14 of 17 higher programmes; the 95% CI included 1 for Acute Internal Medicine, Paediatrics, and Endocrinology and Diabetes Mellitus.

### Ecological association with international applicants

Programmes with a higher percentage of rest-of-world applicants tended to have lower Asian vs White and Black vs White ORs, but none of the correlations was statistically significant (Supplementary Figure S11; Supplementary Table S7). For Asian applicants, Spearman’s correlation was *ρ* = *−*0.45, *p* = 0.224 across 9 entry-level programmes and *ρ* = *−*0.27, *p* = 0.391 across 12 higher programmes. For Black applicants, it was *ρ* = *−*0.30, *p* = 0.433 and *ρ* = *−*0.32, *p* = 0.308, respectively.

### Ethnicity not stated and sensitivity analyses

The percentage of applications with ethnicity not stated ranged from 5.4–10.9% across entry-level programmes and from 3.8–11.8% across higher programmes (Supplementary Figure S14). Applications with ethnicity not stated had lower odds of an offer than those from White applicants in 21 of 23 programmes.

National percentages changed by no more than 0.4 percentage points when suppressed cells were set to 0 or 5 instead of 2.5. Programme-level ORs were robust for Asian applicants (median change 1.8%, largest 13%, no change in statistical significance) and applicants from other ethnic groups. For Black applicants the median change was 11.0%; one comparison (Anaesthetics, higher level) lost statistical significance at one bound. Estimates for the smaller Chinese and Mixed groups were less stable, with a change in statistical significance at one bound for 4 of 17 Chinese and 4 of 17 Mixed comparisons (Supplementary Figure S15; Supplementary Table S8).

## Discussion

Using national data covering five recruitment years, we found that the number of applications to UK specialty training roughly tripled between 2021 and 2025 while the number of offers stayed almost constant, and that the growth came mainly from doctors who qualified outside the UK and EU and from Asian and Black applicants. Across every programme with enough offers to analyse, Asian and Black applicants had substantially lower unadjusted odds of receiving an offer than White applicants, and graduates from outside the UK and EU had substantially lower odds than UK graduates. Results for Chinese applicants were more variable: odds were lower in some entry-level specialties but similar to those of White applicants in most higher specialty programmes. There were clear differences within broad ethnic groups.

The most important caveat is that ethnicity and country of qualification cannot be separated in the published data. Graduates from outside the UK and EU made up 69.4% of applications in 2025 and had much lower offer odds than UK graduates, and they are more likely than UK graduates to be from Asian, Black and other ethnic minority groups. Part, and possibly much, of the difference between broad ethnic groups is therefore likely to reflect country of qualification and the factors that travel with it, such as visa requirements, familiarity with UK portfolios and assessments, and the timing of the expansion in international applications. The within-group results are consistent with this: applicants from ‘other White’ backgrounds, a group likely to include many international graduates, had odds of an offer around a quarter of those of White British applicants at entry level, a difference comparable in size to some of the differences between broad ethnic groups.

Our ecological analysis does not settle the question. Programmes with more rest-of-world applicants tended to have larger ethnic differences, but the correlations were weak to moderate and not statistically significant, and with only 9 entry-level and 12 higher programmes the analysis had little power. Programme-level associations also cannot be used to infer what happens to individual applicants.

Studies that obtained cross-tabulated data have found that the ethnic difference in specialty recruitment success is reduced, but not removed, when country of graduation is taken into account [17, 18]. Differences among UK graduates are arguably the clearest signal of barriers within UK training, and disparities among international graduates raise different questions about induction, support and the conditions under which they apply. Neither can be measured from the data NHS England currently publishes. We recommend that NHS England publish recruitment outcomes cross-tabulated by ethnicity and country of qualification, with appropriate disclosure control.

The Medical Training (Prioritisation) Act 2026 makes this more pressing. Prioritising UK graduates will change who is offered posts, and is likely to narrow the apparent differences between broad ethnic groups in future data, because fewer international graduates will reach the offer stage. It will not, by itself, show whether ethnic minority UK graduates are treated equitably, and without cross-tabulated data a narrowing of the headline gap could be misread as evidence that any ethnic inequity has resolved.

Pakistani and Bangladeshi applicants had lower offer odds than Indian applicants, and Black Caribbean applicants higher odds than Black African applicants, at both training levels and in the largest programmes. These patterns may partly reflect different proportions of UK graduates within each subgroup, which we cannot measure. Whatever their cause, they show that broad categories such as ‘Asian’ or the collective term ‘BAME’ can conceal important differences, and that analyses and interventions should use the most detailed categories the data allow [22, 23].

Specialty recruitment combines several stages, including the Multi-Specialty Recruitment Assessment for many specialties, scored self-assessment and portfolio review, and interviews. Differences can arise at any of them. Ethnic differences in performance in undergraduate and postgraduate assessments are well documented [5, 6]; interviews and other assessments that rely on judgement may be affected by bias, the composition of panels, and differences in communication style [25]; and access to preparation courses, mentorship and portfolio-building opportunities is unequal. Field experiments have shown discrimination at shortlisting in medicine [8] and in the wider labour market [25]. The published data do not include scores at each stage, so we cannot say where the differences arise. Stage-level data, including MSRA scores by ethnicity and country of qualification, would allow this to be examined.

This study uses national data for every specialty and both entry levels across five recruitment years, includes the application, offer and acceptance stages, analyses sub-ethnic groups and country of qualification, and reports the effect of suppressed cells. We verified the published tables against their source and excluded duplicated data.

There are important limitations. First, the ORs are unadjusted, and the data do not allow adjustment for country of qualification, gender, age, socioeconomic background, disability, assessment scores or other characteristics; the results describe differences in outcomes and do not establish their causes. Second, counts are applications rather than applicants, so doctors who apply to several programmes are counted more than once, and the ORs are not independent across programmes. Third, ethnicity offers and acceptances for 2023 were unusable because NHS England published the 2021 tables for that year, so the ethnicity odds ratios are based on 2021, 2022, 2024 and 2025. Fourth, suppression of small counts required imputation; national results were insensitive to this, but estimates for small groups were less stable. Fifth, the 500-offer threshold excluded many smaller specialties, including all programmes in some specialties of interest. Sixth, ethnicity was not stated on 6–8% of applications, and these applications also had lower offer odds than White applicants, so non-response may be associated with outcome. Seventh, we made many comparisons without adjustment for multiplicity, although most of the differences we highlight are very large. Finally, the data contain no information about applicants’ experiences of recruitment, which qualitative research is needed to understand [6].

In conclusion, our findings challenge the assumption that centralised medical recruitment in the UK is entirely meritocratic and free from ethnic bias. While national training pathways offer robust structural protections at initial entry, the persistent disparities observed in speciality-level offer success rates, particularly for Asian and Black candidates and compounded by country of qualification dynamics and international workforce reliance, suggests the recruitment process may benefit reform to improve its processes and remove any unconscious racial bias [17]. Specialities highlighted in our study as having particularly low proportions of non-White doctors may also benefit from wider approaches to promote inclusivity and encourage a diverse pool of applicants to consider joining their speciality.

## Data Availability

The datasets analysed during the current study are available from NHS England at https://medical.hee.nhs.uk/medical-training-recruitment/medical-specialty-training/equality-and-diversity.

## Declarations

### Ethics approval and consent to participate

Not applicable. The data are publicly available aggregate counts, and ethical approval was not required.

### Consent for publication

Not applicable.

### Availability of data and materials

The datasets analysed are published by NHS England at https://medical.hee.nhs.uk/medical-training-recruitment/medical-specialty-training/equality-and-diversity [21]. The analysis code is available from the corresponding author on request.

## Authors’ contributions

All authors conceived the study, analysed and interpreted the findings, and read and approved the final manuscript.

## Acknowledgements

Not applicable.

## Supplementary information

Supplementary Methods, Supplementary Figures S1–S15 and Supplementary Tables S1–S8 are provided in a separate file.

