## Supplementary Materials for "Recruitment Success Among Ethnic Minority Doctors and International Medical Graduates in UK Medical Specialty Recruitment: A Repeated Cross-Sectional Analysis of NHS England Data (2021–2025)"

**2 Supplementary Figures** 3

**3 Supplementary Tables** 12

### Supplementary Methods

#### Source data

NHS England publishes annual equality and diversity data for national specialty training recruitment, with separate web pages for each protected characteristic and recruitment year. Each page contains four tables, by programme and entry level: applications made, appointable applicants, offers made, and offers accepted. We used the ethnic origin and country of qualification pages for 2021 to 2025. The appointable-applicants stage was not analysed.

#### Data verification and corrections

Every table used was checked against the published web pages; snapshots of the pages were saved on 10 November 2026. Two problems were found.

**Ethnicity offers and acceptances, 2023.** On the 2023 ethnic origin page, the offers made and offers accepted tables are identical, cell for cell, to the corresponding 2021 tables. They have the 2021 programme list (58 rows, against 69 rows in the 2023 applications table) and the 2021 category labels (for example 'Asian/Asian British Indian' rather than the 2023 'Asian or Asian British - Indian'). The 2023 applications table is specific to 2023. We therefore excluded 2023 ethnicity offers and acceptances. Because odds ratios are calculated only from programme-years for which both applications and offers are available, 2023 contributes to no ethnicity odds ratio, and the ethnicity odds ratios are based on the

2021, 2022, 2024 and 2025 recruitment years. Applications for 2023 are included in descriptive results. The country of qualification tables for 2023 are not affected.

**Country of qualification, 2021–2023.** An earlier compilation of these years had assigned each table to the following stage, so that appointable applicants were recorded as offers and offers as acceptances; 2024 offers had also been omitted. All country of qualification data were re-extracted from the published tables. After correction, offers by ethnicity and by country of qualification agree closely in every year (for example, 16,988 and 16,771 offers in 2021).

### Programme harmonisation and training level

Programme titles were standardised across years. Acute Care Common Stem routes were combined with their parent specialty (Emergency Medicine, Anaesthetics, and Internal Medicine Training). Other variant titles were mapped to a single name (for example ‘Oral and Maxillo-facial Surgery’ and ‘Oral and Maxillofacial Surgery’; ‘Core Psychiatry Training’ and ‘Core Psychiatry’). Each programme was assigned to entry-level training (published level containing 1 or 2, CT, or GP) or higher specialty training (level 3 or above).

### Suppressed cells

Counts below five are published as ‘<5’. These were set to 2.5 in the primary analysis and to 0 and 5 in sensitivity analyses (Supplementary Figure S15, Supplementary Table S8). The 500-offer threshold was applied separately under each value, so a small number of comparisons meet the threshold under one value but not another; these are omitted from the sensitivity comparison.

### Odds ratios

For each programme, applications ($n$) and offers ($k$) were summed across recruitment years for the group of interest and the reference group. The odds ratio was

$$\mathrm{OR} = \frac{k_1/(n_1 - k_1)}{k_0/(n_0 - k_0)}, \qquad \mathrm{SE}(\log \mathrm{OR}) = \sqrt{\tfrac{1}{k_1} + \tfrac{1}{n_1 - k_1} + \tfrac{1}{k_0} + \tfrac{1}{n_0 - k_0}},$$

with 0.5 added to each cell only when a cell was zero. Programmes were included if the cumulative offers to applicants of known ethnicity (or, for country of qualification, to all applicants) were at least 500. Within-group odds ratios were pooled across included programmes by summing offers and applications (crude); Mantel–Haenszel odds ratios stratified by programme, with the Robins–Breslow–Greenland variance, are given for comparison (Supplementary Table S5).

### Supplementary Figures

**Figure S1** Sub-ethnic composition of applications by recruitment year, 2021–2025. Labels show percentages of at least 4%. . . . . 4

**Figure S2** Sub-ethnic composition of applications by specialty: (a) entry-level (ST1/CT1) and (b) higher specialty (ST3+) programmes, 2021–2025. Programmes with fewer than 500 cumulative applications are not shown. 4

**Figure S3** Ethnic composition of offers by specialty: (a) entry-level and (b) higher specialty programmes, 2021, 2022, 2024 and 2025. Programmes with fewer than 500 cumulative offers are not shown. . . . . 5

**Figure S4** Sub-ethnic composition of offers by specialty: (a) entry-level and (b) higher specialty programmes, 2021, 2022, 2024 and 2025. Programmes with fewer than 500 cumulative offers are not shown. . . . . 5

**Figure S5** Ethnic composition of accepted offers by specialty: (a) entry-level and (b) higher specialty programmes, 2021, 2022, 2024 and 2025. Programmes with fewer than 500 cumulative acceptances are not shown. 5

**Figure S6** Sub-ethnic composition of accepted offers by specialty: (a) entry-level and (b) higher specialty programmes, 2021, 2022, 2024 and 2025. Programmes with fewer than 500 cumulative acceptances are not shown. 6

**Figure S7** Country of primary medical qualification of offers by specialty: (a) entry-level and (b) higher specialty programmes, 2021–2025. Programmes with fewer than 500 cumulative offers are not shown. . . . . 6

**Figure S8** Country of primary medical qualification of accepted offers by specialty: (a) entry-level and (b) higher specialty programmes, 2021–2025. Programmes with fewer than 500 cumulative acceptances are not shown. . . . . 6

**Figure S9** Odds of receiving an offer within broad ethnic groups, by training level. Each subgroup is compared with the largest subgroup in its broad group (Indian, Black African or White British). Crude odds ratios pooled across programmes with at least 500 cumulative offers, with 95% CIs on a log scale. Filled points: 95% CI excludes 1. Mantel–Haenszel estimates are in Supplementary Table S5. . . . . 7

**Figure S10** Odds of receiving an offer for (a, b) EU/EEA graduates and (c, d) graduates from the rest of the world compared with UK graduates, by programme, for entry-level (left) and higher specialty (right) programmes. Odds ratios with 95% CIs on a log scale; filled points: 95% CI excludes 1. Programmes with at least 500 cumulative offers, 2021–2025. . . . . 8

**Figure S11** Programme-level association between the percentage of applications from graduates outside the UK and EU and the offer odds ratio for (a, b) Asian and (c, d) Black applicants compared with White applicants, for entry-level (left) and higher specialty (right) programmes. Each point is a programme with at least 500 cumulative offers; the two lowest and two highest points are labelled, and all values are listed in Supplementary Table S7. Dashed lines: least-squares fit of log odds ratio on percentage, shown as a visual guide. Horizontal line: OR=1. Spearman's $\rho$ and $p$ values are shown above each panel. . . . . 9

**Figure S12** Odds of receiving an offer for applicants from (a, b) Mixed and (c, d) other ethnic groups compared with White applicants, by programme. Odds ratios with 95% CIs on a log scale; filled points: 95% CI excludes 1. Programmes with at least 500 cumulative offers in 2021, 2022, 2024 and 2025. . . . . 10

**Figure S13** Share of each ethnic group at the application, offer and acceptance stages, for (a) entry-level and (b) higher specialty programmes. Calculated over programme-years with data at all three stages (2021, 2022, 2024 and 2025). . . . . 10

**Figure S14** Percentage of applications with ethnicity not stated, by programme, for (a) entry-level and (b) higher specialty programmes, 2021–2025. Dashed line: percentage across all programmes. Programmes with fewer than 500 cumulative applications are not shown. . . . . 11

**Figure S15** Sensitivity of offer odds ratios to the value used for suppressed cells, for (a) Asian, (b) Black and (c) Chinese applicants compared with White applicants. Each point is a programme; the x-axis shows the primary estimate (suppressed cells set to 2.5) and the y-axis the estimate with suppressed cells set to 0 (circles) or 5 (squares). Points on the diagonal are unchanged. Log scales. . . . . 11

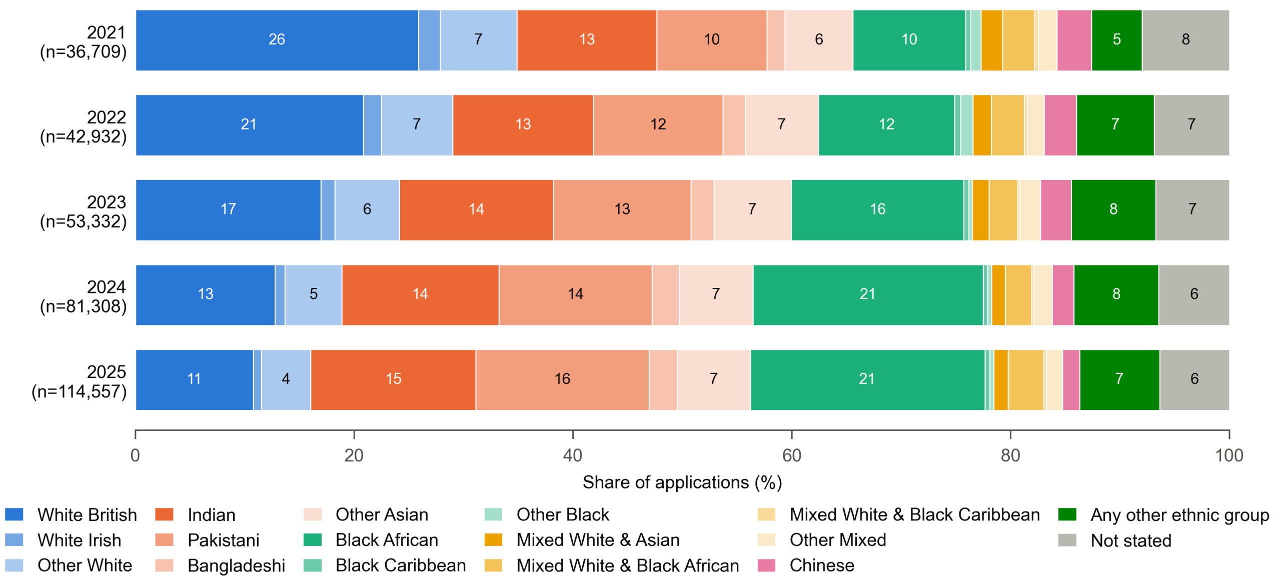

**Supplementary Figure S1.** Sub-ethnic composition of applications by recruitment year, 2021–2025. Labels show percentages of at least 4%.

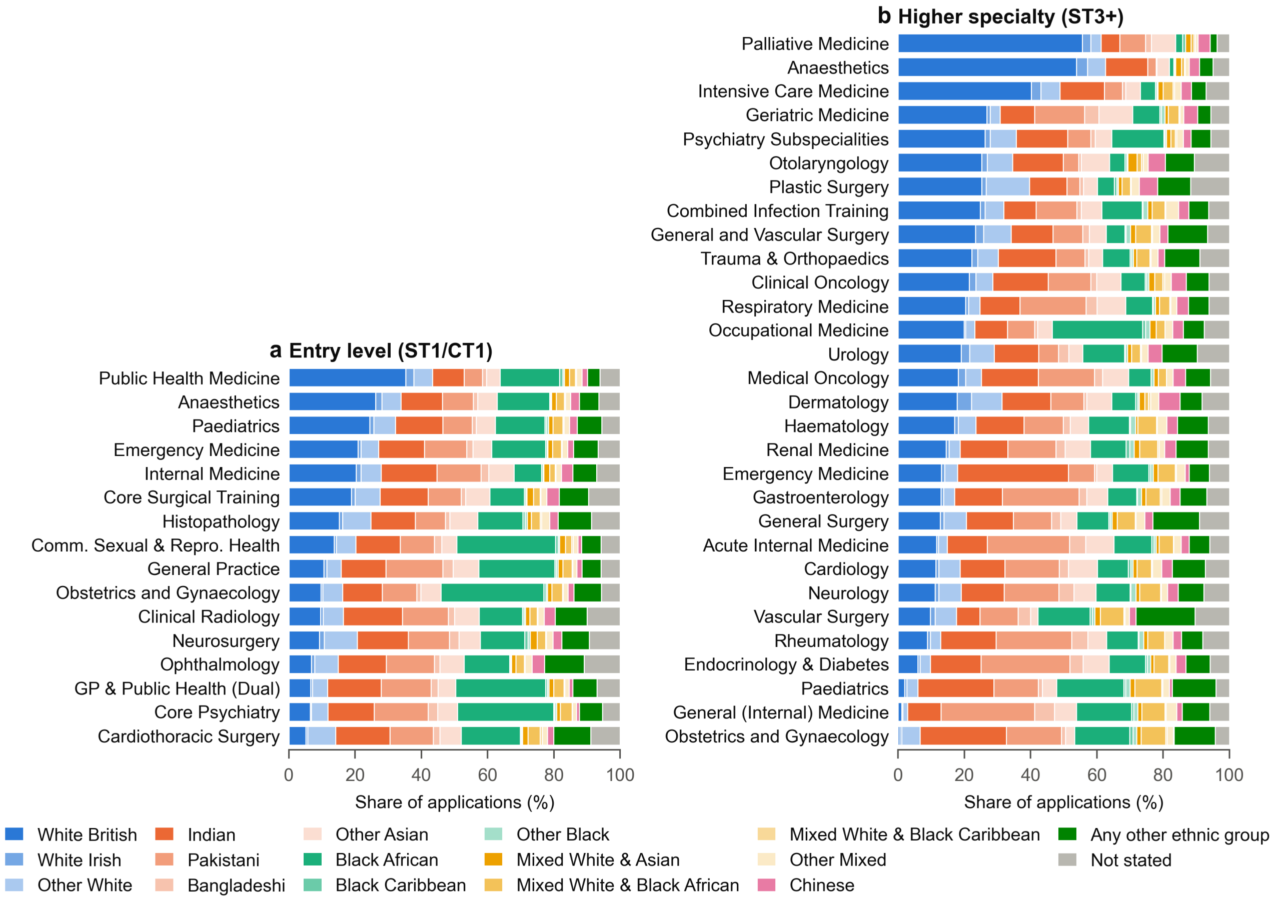

**Supplementary Figure S2.** Sub-ethnic composition of applications by specialty: (a) entry-level (ST1/CT1) and (b) higher specialty (ST3+) programmes, 2021–2025. Programmes with fewer than 500 cumulative applications are not shown.

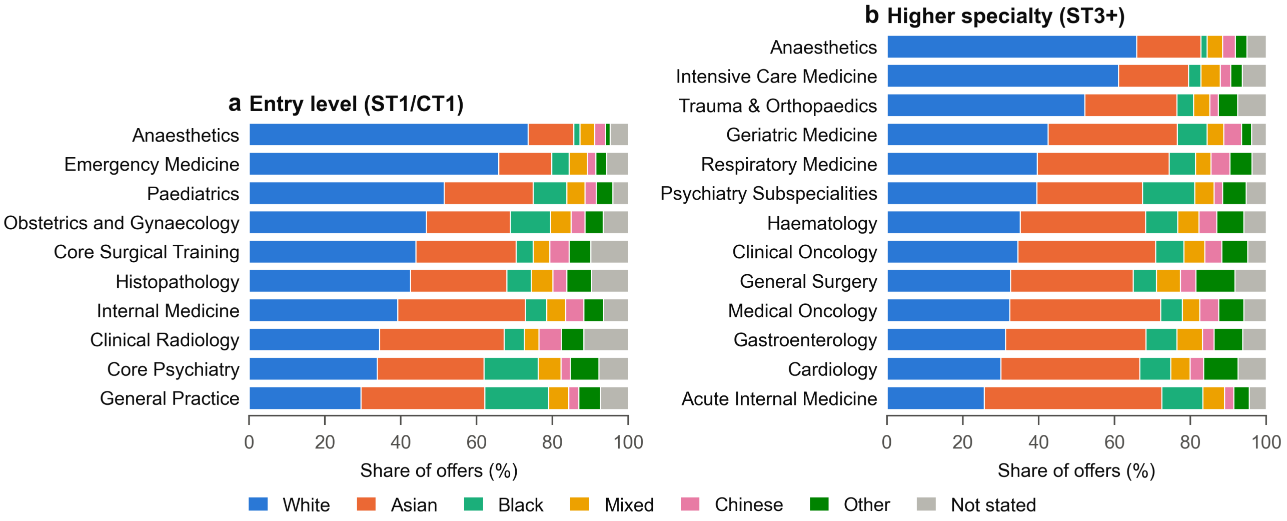

**Supplementary Figure S3.** Ethnic composition of offers by specialty: (a) entry-level and (b) higher specialty programmes, 2021, 2022, 2024 and 2025. Programmes with fewer than 500 cumulative offers are not shown.

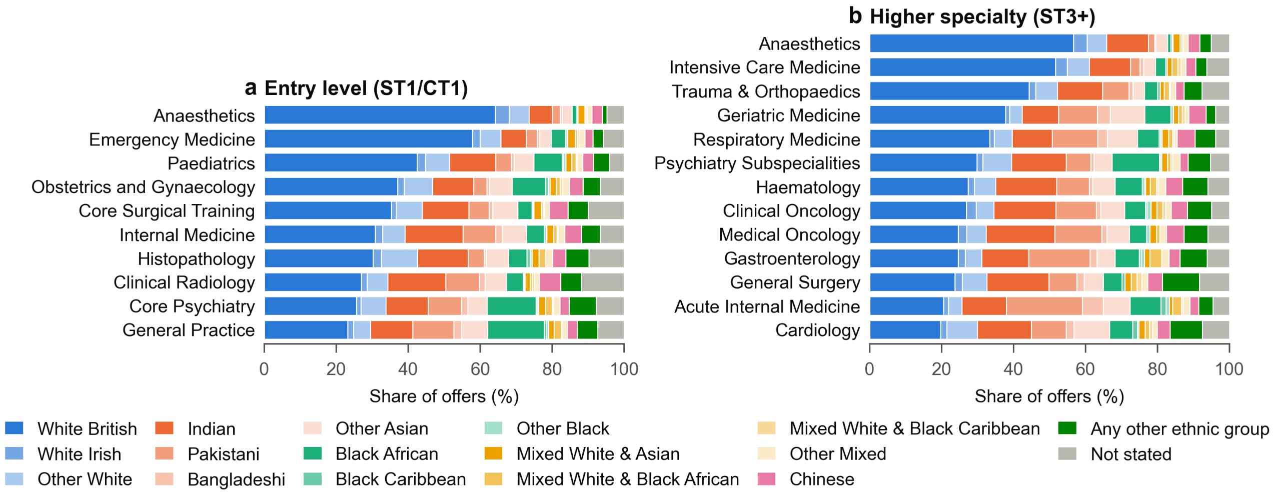

**Supplementary Figure S4.** Sub-ethnic composition of offers by specialty: (a) entry-level and (b) higher specialty programmes, 2021, 2022, 2024 and 2025. Programmes with fewer than 500 cumulative offers are not shown.

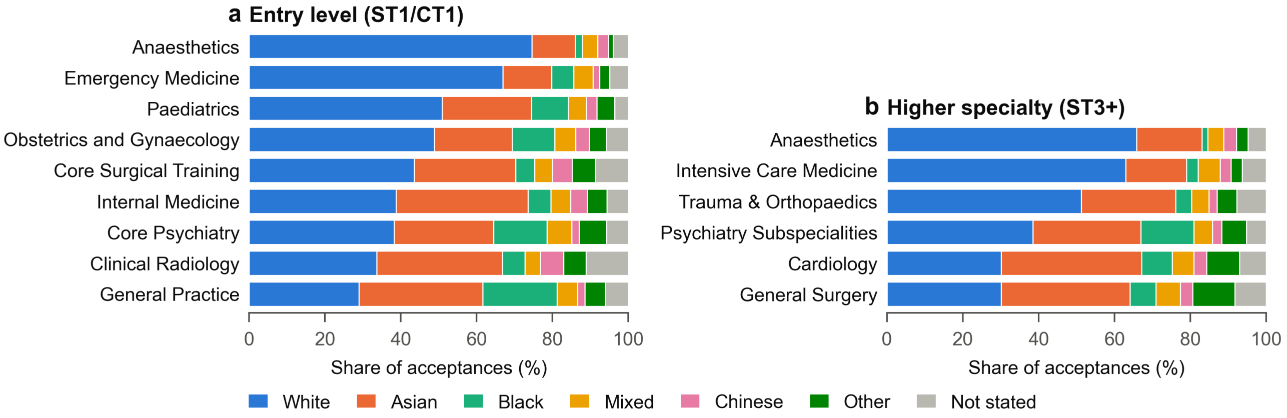

**Supplementary Figure S5.** Ethnic composition of accepted offers by specialty: (a) entry-level and (b) higher specialty programmes, 2021, 2022, 2024 and 2025. Programmes with fewer than 500 cumulative acceptances are not shown.

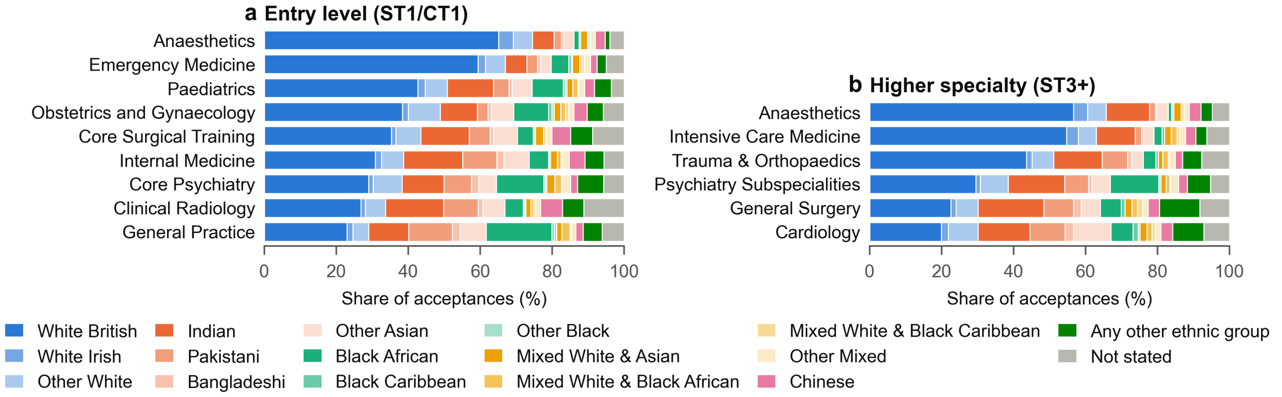

**Supplementary Figure S6.** Sub-ethnic composition of accepted offers by specialty: (a) entry-level and (b) higher specialty programmes, 2021, 2022, 2024 and 2025. Programmes with fewer than 500 cumulative acceptances are not shown.

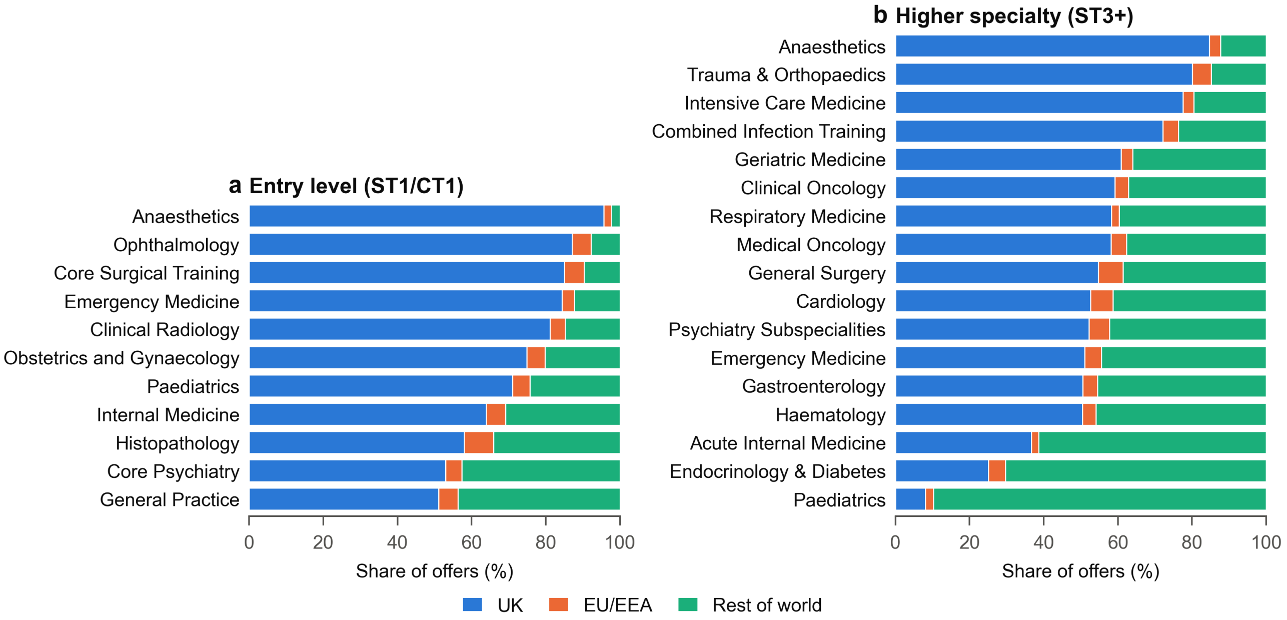

**Supplementary Figure S7.** Country of primary medical qualification of offers by specialty: (a) entry-level and (b) higher specialty programmes, 2021–2025. Programmes with fewer than 500 cumulative offers are not shown.

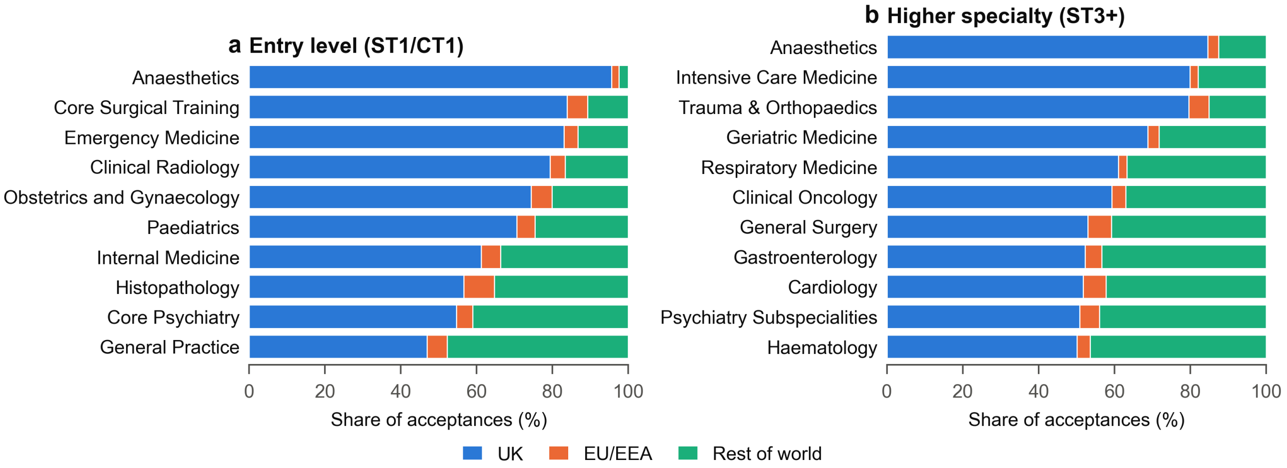

**Supplementary Figure S8.** Country of primary medical qualification of accepted offers by specialty: (a) entry-level and (b) higher specialty programmes, 2021–2025. Programmes with fewer than 500 cumulative acceptances are not shown.

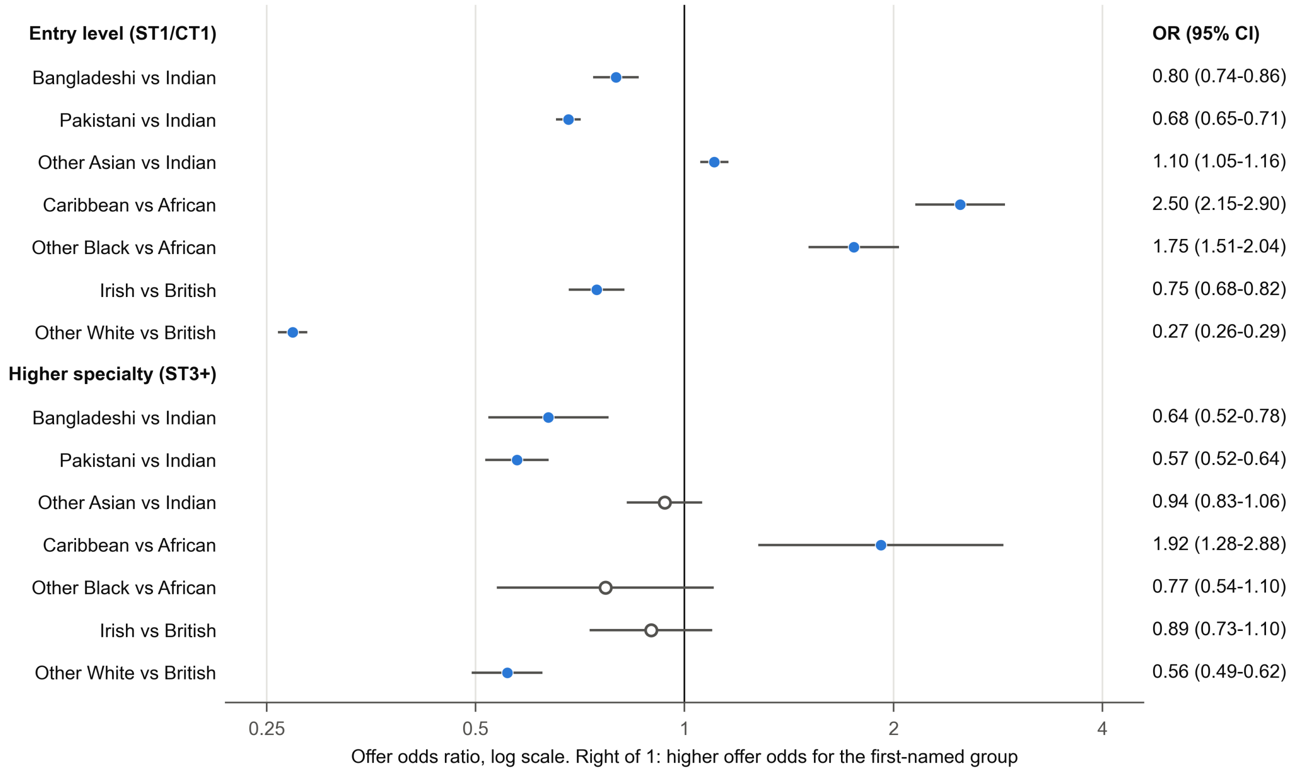

**Supplementary Figure S9.** Odds of receiving an offer within broad ethnic groups, by training level. Each subgroup is compared with the largest subgroup in its broad group (Indian, Black African or White British). Crude odds ratios pooled across programmes with at least 500 cumulative offers, with 95% CIs on a log scale. Filled points: 95% CI excludes 1. Mantel–Haenszel estimates are in Supplementary Table S5.

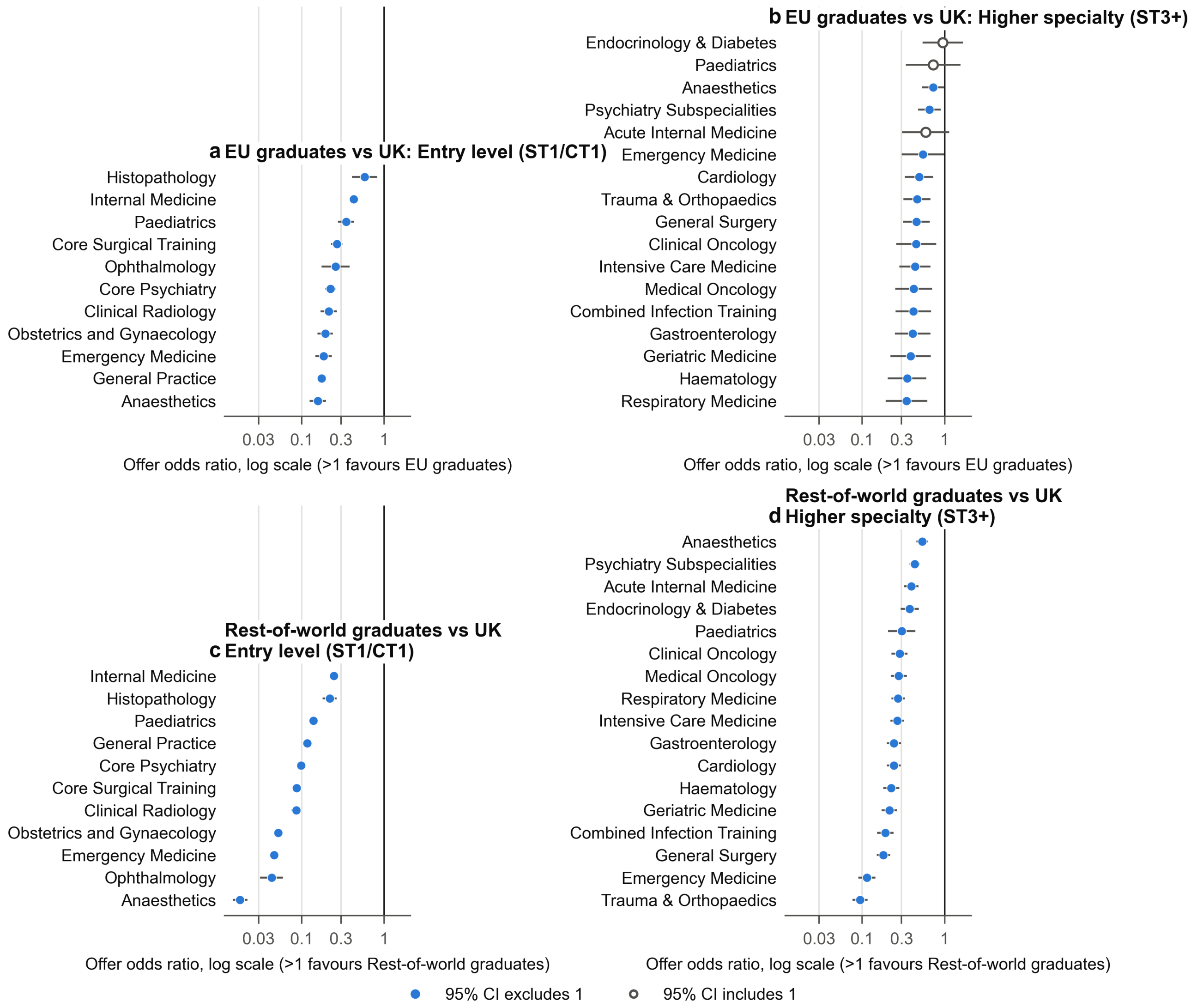

**Supplementary Figure S10.** Odds of receiving an offer for (a, b) EU/EEA graduates and (c, d) graduates from the rest of the world compared with UK graduates, by programme, for entry-level (left) and higher specialty (right) programmes. Odds ratios with 95% CIs on a log scale; filled points: 95% CI excludes 1. Programmes with at least 500 cumulative offers, 2021–2025.

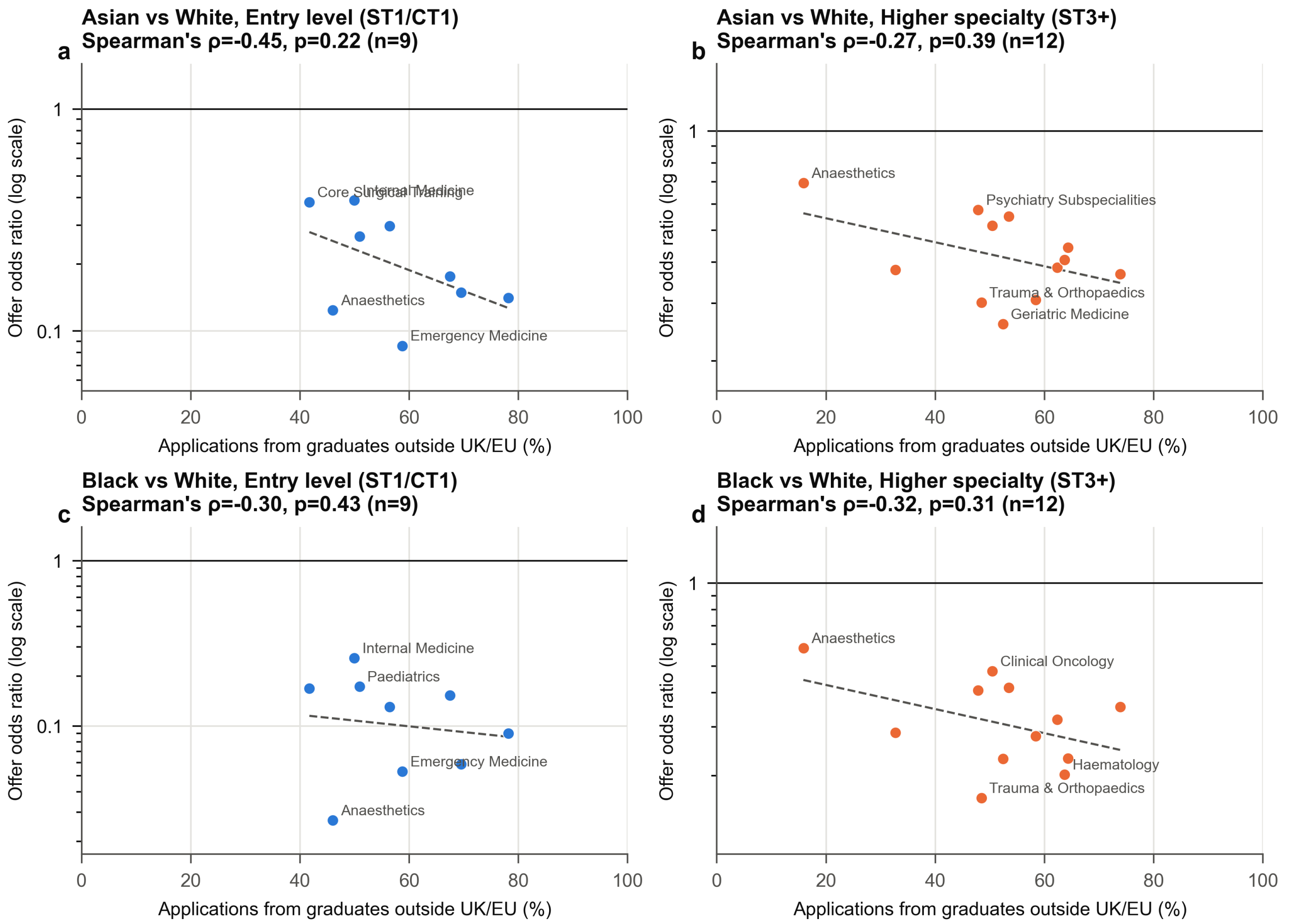

**Supplementary Figure S11.** Programme-level association between the percentage of applications from graduates outside the UK and EU and the offer odds ratio for (a, b) Asian and (c, d) Black applicants compared with White applicants, for entry-level (left) and higher specialty (right) programmes. Each point is a programme with at least 500 cumulative offers; the two lowest and two highest points are labelled, and all values are listed in Supplementary Table S7. Dashed lines: least-squares fit of log odds ratio on percentage, shown as a visual guide. Horizontal line: OR=1. Spearman's $\rho$ and $p$ values are shown above each panel.

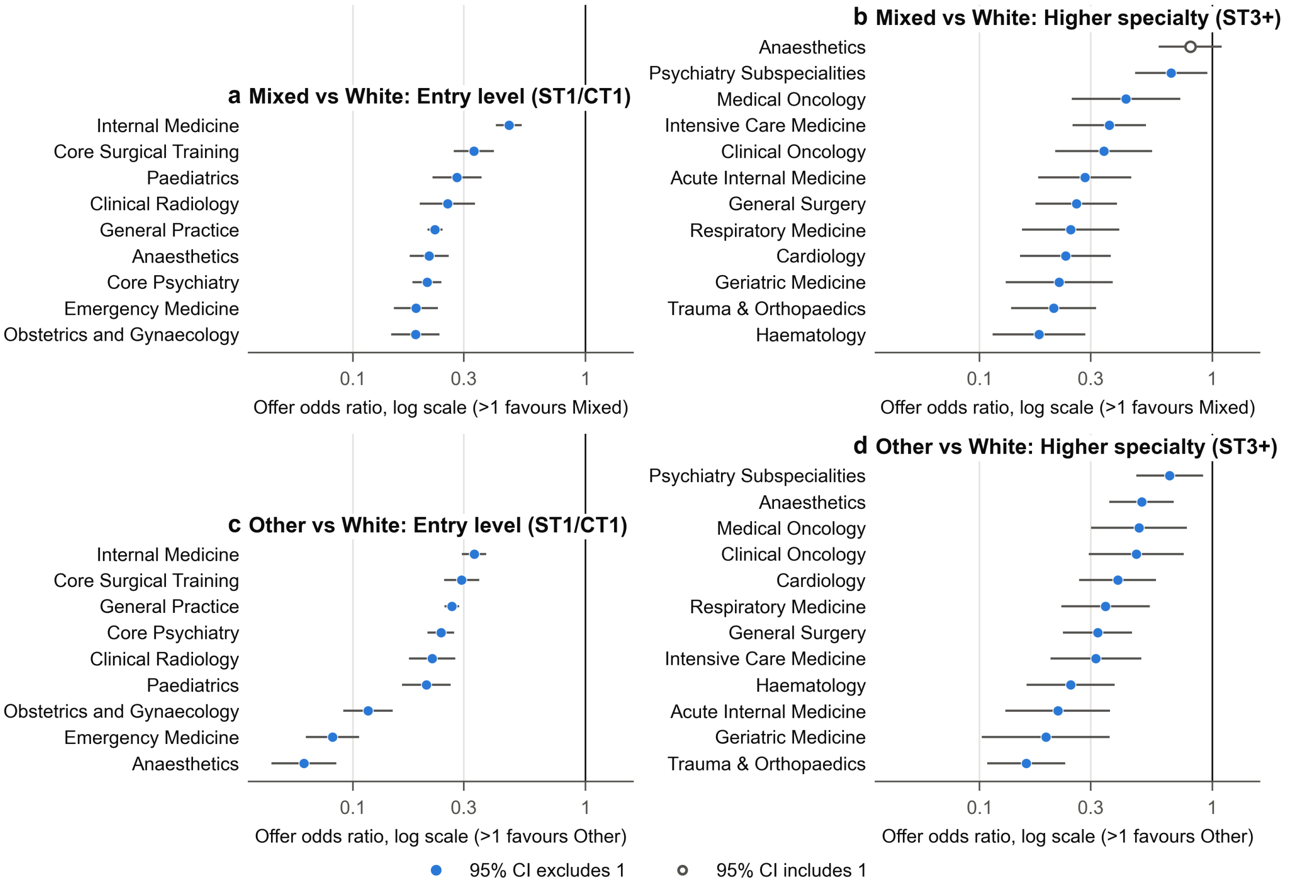

**Supplementary Figure S12.** Odds of receiving an offer for applicants from (a, b) Mixed and (c, d) other ethnic groups compared with White applicants, by programme. Odds ratios with 95% CIs on a log scale; filled points: 95% CI excludes 1. Programmes with at least 500 cumulative offers in 2021, 2022, 2024 and 2025.

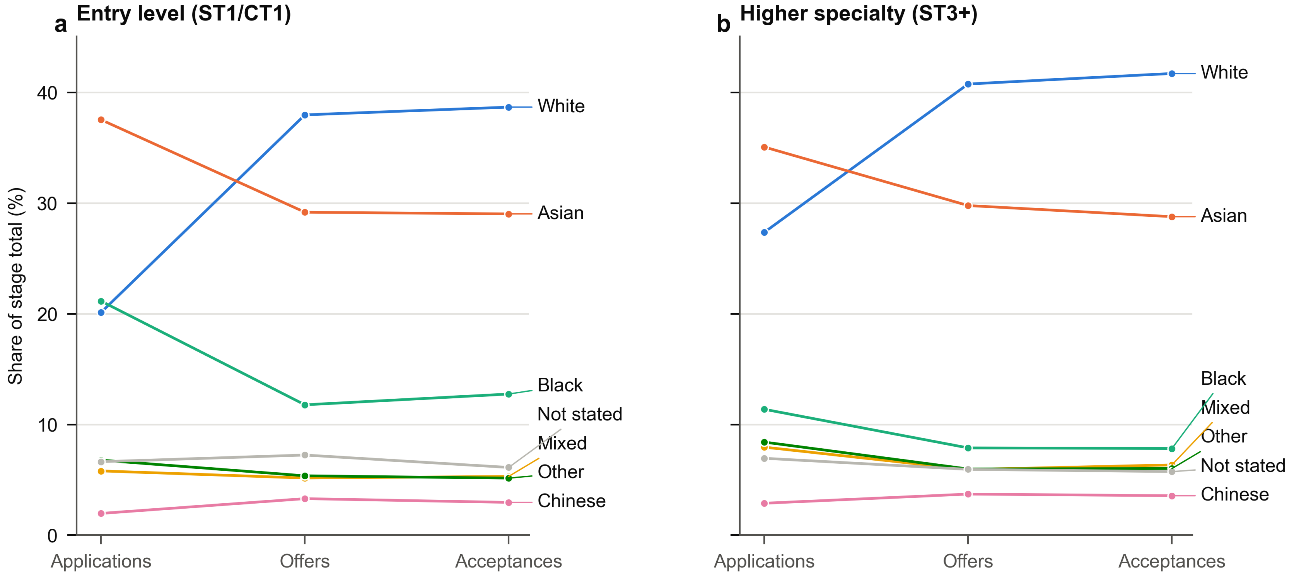

**Supplementary Figure S13.** Share of each ethnic group at the application, offer and acceptance stages, for (a) entry-level and (b) higher specialty programmes. Calculated over programme-years with data at all three stages (2021, 2022, 2024 and 2025).

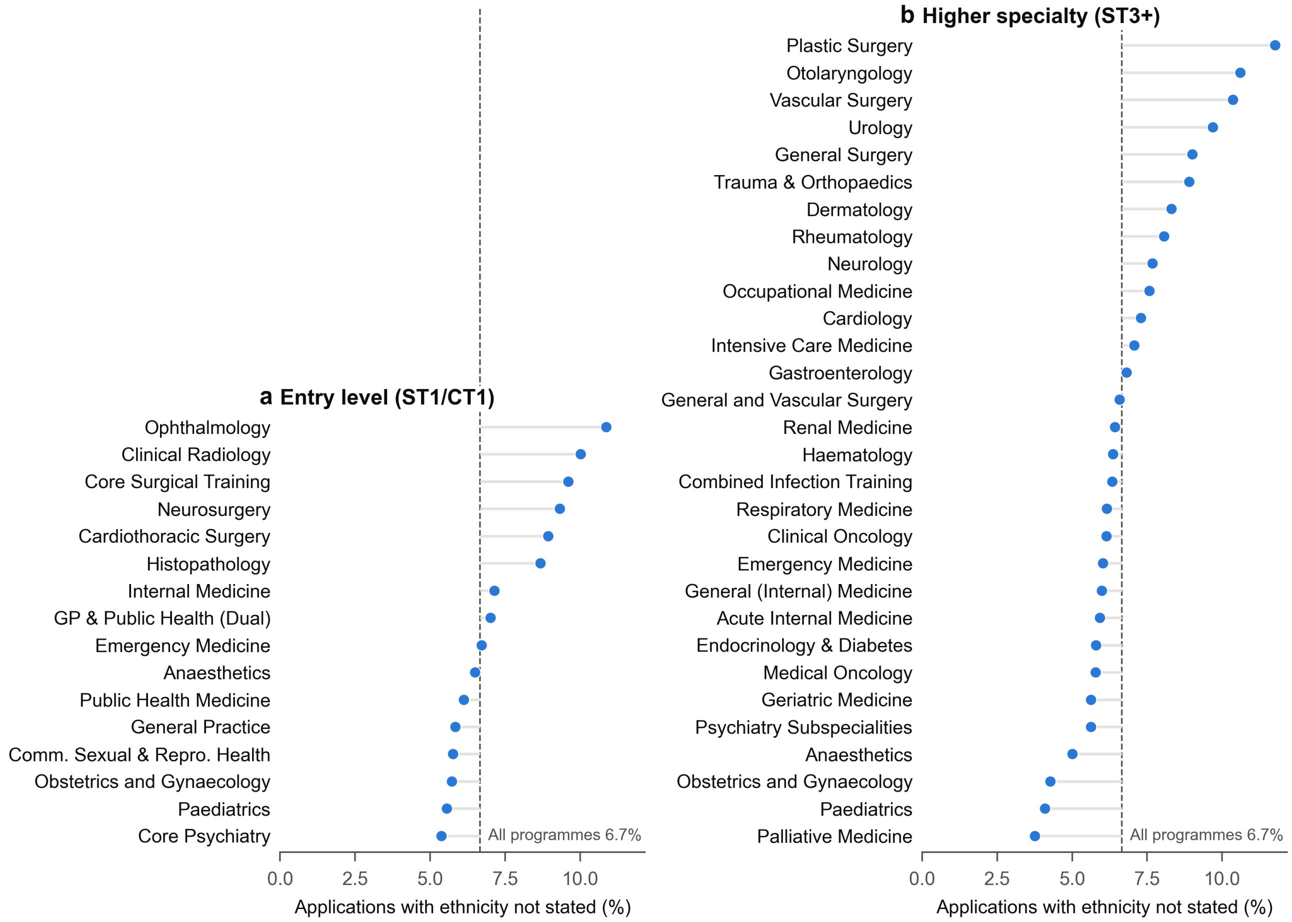

**Supplementary Figure S14.** Percentage of applications with ethnicity not stated, by programme, for (a) entry-level and (b) higher specialty programmes, 2021–2025. Dashed line: percentage across all programmes. Programmes with fewer than 500 cumulative applications are not shown.

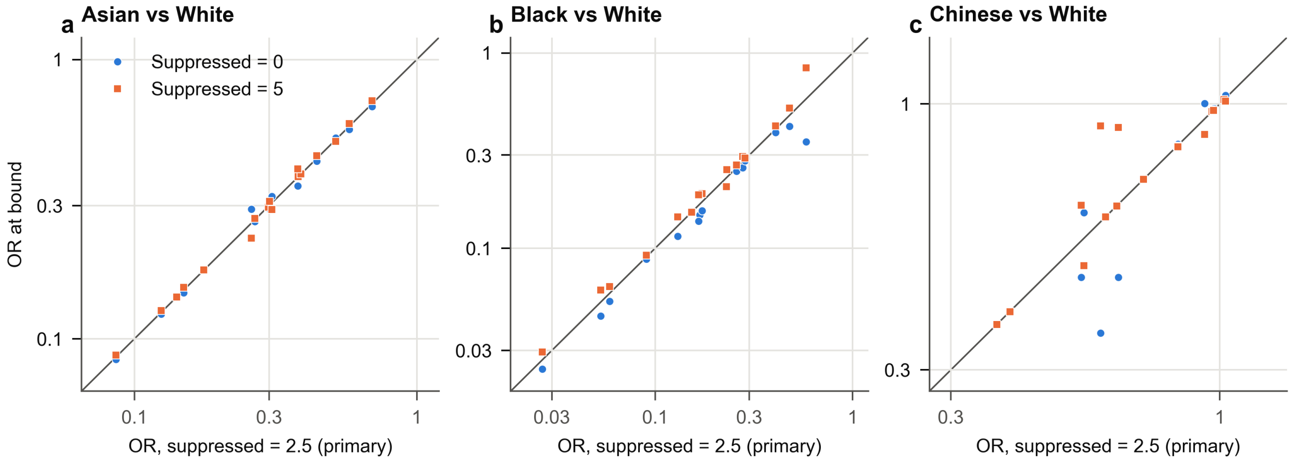

**Supplementary Figure S15.** Sensitivity of offer odds ratios to the value used for suppressed cells, for (a) Asian, (b) Black and (c) Chinese applicants compared with White applicants. Each point is a programme; the x-axis shows the primary estimate (suppressed cells set to 2.5) and the y-axis the estimate with suppressed cells set to 0 (circles) or 5 (squares). Points on the diagonal are unchanged. Log scales.

### Supplementary Tables

**Table S1** Applications, offers and acceptances by broad ethnic group and recruitment year, n (%). 2023 offers and acceptances are not shown because the tables NHS England published for 2023 are copies of the 2021 tables (Supplementary Methods). . . . 13

**Table S2** Applications, offers and acceptances by country of primary medical qualification and recruitment year, n (%). . . . 14

**Table S3** Offer odds ratios (95% CI) for each broad ethnic group compared with White applicants, by programme. Programmes with at least 500 cumulative offers, 2021, 2022, 2024 and 2025. . . . 15

**Table S4** Offer odds ratios (95% CI) by country of primary medical qualification, compared with UK graduates. Programmes with at least 500 cumulative offers, 2021–2025. . . . 16

**Table S5** Offer odds ratios within broad ethnic groups, pooled over programmes with at least 500 cumulative offers. Crude: offers and applications summed across programmes (primary analysis). MH: Mantel–Haenszel odds ratio stratified by programme. . . . 17

**Table S6** Within-group offer odds ratios for the three largest entry-level programmes. Target and reference are the first- and second-named groups in each comparison. . . . 17

**Table S7** Programme-level data for the ecological analysis: percentage of applications from graduates outside the UK and EU (ROW, 2021–2025) and offer odds ratios for Asian and Black applicants compared with White applicants. . . . 18

**Table S8** Offer odds ratios with suppressed cells (fewer than five) set to 0, 2.5 (primary) and 5. Comparisons that fall below the 500-offer threshold under one of the bounds are omitted. . . . 19

**Supplementary Table S1.** Applications, offers and acceptances by broad ethnic group and recruitment year, n (%). 2023 offers and acceptances are not shown because the tables NHS England published for 2023 are copies of the 2021 tables (Supplementary Methods).

| Stage | Year | White | Asian | Black | Mixed | Chinese | Other | Not stated | Total |
|---|---|---|---|---|---|---|---|---|---|
| Applications | 2021 | 12,797 (34.9) | 11,278 (30.7) | 4,300 (11.7) | 2,543 (6.9) | 1,162 (3.2) | 1,701 (4.6) | 2,928 (8.0) | 36,709 |
| Applications | 2022 | 12,452 (29.0) | 14,349 (33.4) | 6,060 (14.1) | 2,801 (6.5) | 1,270 (3.0) | 3,054 (7.1) | 2,946 (6.9) | 42,932 |
| Applications | 2023 | 12,876 (24.1) | 19,094 (35.8) | 8,825 (16.5) | 3,326 (6.2) | 1,502 (2.8) | 4,122 (7.7) | 3,588 (6.7) | 53,332 |
| Applications | 2024 | 15,337 (18.9) | 30,562 (37.6) | 17,742 (21.8) | 4,492 (5.5) | 1,602 (2.0) | 6,322 (7.8) | 5,250 (6.5) | 81,308 |
| Applications | 2025 | 18,332 (16.0) | 46,052 (40.2) | 25,494 (22.3) | 7,179 (6.3) | 1,818 (1.6) | 8,376 (7.3) | 7,306 (6.4) | 114,557 |
| Offers | 2021 | 7,291 (42.9) | 4,604 (27.1) | 1,606 (9.5) | 961 (5.7) | 627 (3.7) | 738 (4.3) | 1,160 (6.8) | 16,988 |
| Offers | 2022 | 7,130 (38.3) | 5,506 (29.6) | 2,152 (11.6) | 988 (5.3) | 644 (3.5) | 1,026 (5.5) | 1,148 (6.2) | 18,594 |
| Offers | 2023 | – | – | – | – | – | – | – | – |
| Offers | 2024 | 7,282 (36.9) | 5,884 (29.9) | 2,258 (11.5) | 1,023 (5.2) | 665 (3.4) | 1,194 (6.1) | 1,405 (7.1) | 19,710 |
| Offers | 2025 | 7,217 (36.9) | 5,953 (30.4) | 2,109 (10.8) | 1,027 (5.3) | 606 (3.1) | 1,164 (6.0) | 1,475 (7.5) | 19,551 |
| Acceptances | 2021 | 5,516 (43.7) | 3,320 (26.3) | 1,262 (10.0) | 752 (6.0) | 420 (3.3) | 532 (4.2) | 806 (6.4) | 12,608 |
| Acceptances | 2022 | 5,242 (39.5) | 3,829 (28.9) | 1,564 (11.8) | 733 (5.5) | 436 (3.3) | 702 (5.3) | 750 (5.7) | 13,256 |
| Acceptances | 2023 | – | – | – | – | – | – | – | – |
| Acceptances | 2024 | 5,366 (38.0) | 4,202 (29.7) | 1,734 (12.3) | 752 (5.3) | 423 (3.0) | 834 (5.9) | 819 (5.8) | 14,132 |
| Acceptances | 2025 | 5,457 (37.2) | 4,482 (30.6) | 1,704 (11.6) | 814 (5.5) | 425 (2.9) | 872 (5.9) | 914 (6.2) | 14,668 |

**Supplementary Table S2.** Applications, offers and acceptances by country of primary medical qualification and recruitment year, n (%).

| Stage | Year | UK | EU/EEA | Rest of world | Total |
|---|---|---|---|---|---|
| Applications | 2021 | 18,200 (49.8) | 2,312 (6.3) | 16,066 (43.9) | 36,578 |
| Applications | 2022 | 17,916 (42.3) | 2,902 (6.9) | 21,490 (50.8) | 42,306 |
| Applications | 2023 | 19,570 (37.1) | 3,303 (6.3) | 29,852 (56.6) | 52,726 |
| Applications | 2024 | 24,080 (29.9) | 5,236 (6.5) | 51,260 (63.6) | 80,576 |
| Applications | 2025 | 31,252 (27.3) | 3,814 (3.3) | 79,406 (69.4) | 114,472 |
| Offers | 2021 | 11,024 (65.7) | 812 (4.8) | 4,936 (29.4) | 16,771 |
| Offers | 2022 | 10,848 (59.0) | 982 (5.3) | 6,560 (35.7) | 18,391 |
| Offers | 2023 | 11,806 (59.6) | 909 (4.6) | 7,104 (35.8) | 19,818 |
| Offers | 2024 | 12,260 (62.9) | 870 (4.5) | 6,370 (32.7) | 19,499 |
| Offers | 2025 | 11,410 (59.0) | 898 (4.6) | 7,044 (36.4) | 19,352 |
| Acceptances | 2021 | 8,040 (64.9) | 600 (4.8) | 3,754 (30.3) | 12,394 |
| Acceptances | 2022 | 7,678 (58.9) | 714 (5.5) | 4,634 (35.6) | 13,027 |
| Acceptances | 2023 | 8,123 (58.9) | 638 (4.6) | 5,029 (36.5) | 13,790 |
| Acceptances | 2024 | 8,543 (61.4) | 642 (4.6) | 4,724 (34.0) | 13,908 |
| Acceptances | 2025 | 8,270 (57.2) | 637 (4.4) | 5,546 (38.4) | 14,453 |

**Supplementary Table S3.** Offer odds ratios (95% CI) for each broad ethnic group compared with White applicants, by programme. Programmes with at least 500 cumulative offers, 2021, 2022, 2024 and 2025.

| Programme | Level | Asian | Black | Chinese | Mixed | Other |
|---|---|---:|---:|---:|---:|---:|
| Anaesthetics | ST1/CT1 | 0.12 (0.11–0.14) | 0.027 (0.020–0.036) | 0.39 (0.31–0.49) | 0.21 (0.18–0.26) | 0.062 (0.045–0.085) |
| Clinical Radiology | ST1/CT1 | 0.30 (0.26–0.34) | 0.13 (0.10–0.16) | 0.83 (0.65–1.06) | 0.26 (0.19–0.34) | 0.22 (0.17–0.28) |
| Core Psychiatry | ST1/CT1 | 0.14 (0.13–0.15) | 0.090 (0.081–0.100) | 0.97 (0.75–1.24) | 0.21 (0.18–0.24) | 0.24 (0.21–0.27) |
| Core Surgical Training | ST1/CT1 | 0.38 (0.34–0.42) | 0.17 (0.14–0.20) | 0.83 (0.68–1.01) | 0.33 (0.27–0.40) | 0.29 (0.25–0.35) |
| Emergency Medicine | ST1/CT1 | 0.086 (0.075–0.098) | 0.053 (0.043–0.066) | 0.37 (0.26–0.52) | 0.19 (0.15–0.23) | 0.082 (0.063–0.106) |
| General Practice | ST1/CT1 | 0.18 (0.17–0.18) | 0.15 (0.15–0.16) | 1.02 (0.89–1.17) | 0.23 (0.21–0.24) | 0.27 (0.25–0.29) |
| Internal Medicine | ST1/CT1 | 0.39 (0.36–0.41) | 0.26 (0.23–0.29) | 0.97 (0.84–1.13) | 0.47 (0.41–0.53) | 0.33 (0.29–0.37) |
| Obstetrics and Gynaecology | ST1/CT1 | 0.15 (0.13–0.17) | 0.059 (0.049–0.070) | 0.60 (0.44–0.82) | 0.19 (0.15–0.24) | 0.116 (0.091–0.148) |
| Paediatrics | ST1/CT1 | 0.27 (0.23–0.30) | 0.17 (0.14–0.21) | 0.63 (0.45–0.89) | 0.28 (0.22–0.36) | 0.21 (0.16–0.26) |
| Acute Internal Medicine | ST3+ | 0.37 (0.28–0.48) | 0.35 (0.24–0.52) | 0.59 (0.29–1.20) | 0.28 (0.18–0.45) | 0.22 (0.13–0.36) |
| Anaesthetics | ST3+ | 0.70 (0.59–0.82) | 0.58 (0.37–0.92) | 0.97 (0.68–1.40) | 0.80 (0.59–1.10) | 0.50 (0.36–0.68) |
| Cardiology | ST3+ | 0.38 (0.30–0.50) | 0.32 (0.22–0.47) | 0.72 (0.39–1.31) | 0.23 (0.15–0.37) | 0.39 (0.27–0.57) |
| Clinical Oncology | ST3+ | 0.52 (0.39–0.69) | 0.48 (0.30–0.75) | 0.63 (0.35–1.15) | 0.34 (0.21–0.55) | 0.47 (0.29–0.75) |
| General Surgery | ST3+ | 0.44 (0.34–0.57) | 0.23 (0.15–0.35) | 1.03 (0.57–1.84) | 0.26 (0.17–0.39) | 0.32 (0.23–0.45) |
| Geriatric Medicine | ST3+ | 0.26 (0.19–0.35) | 0.23 (0.15–0.35) | 0.54 (0.29–1.01) | 0.22 (0.13–0.37) | 0.19 (0.10–0.36) |
| Haematology | ST3+ | 0.41 (0.30–0.55) | 0.20 (0.13–0.30) | 0.93 (0.48–1.81) | 0.18 (0.11–0.29) | 0.25 (0.16–0.38) |
| Intensive Care Medicine | ST3+ | 0.38 (0.30–0.47) | 0.29 (0.19–0.44) | 0.54 (0.32–0.90) | 0.36 (0.25–0.52) | 0.32 (0.20–0.50) |
| Medical Oncology | ST3+ | 0.55 (0.41–0.73) | 0.42 (0.26–0.68) | 1.22 (0.64–2.33) | 0.43 (0.25–0.73) | 0.48 (0.30–0.78) |
| Psychiatry Subspecialities | ST3+ | 0.58 (0.47–0.70) | 0.41 (0.33–0.51) | 0.71 (0.42–1.20) | 0.67 (0.47–0.95) | 0.66 (0.47–0.91) |
| Respiratory Medicine | ST3+ | 0.31 (0.24–0.39) | 0.28 (0.19–0.41) | 0.93 (0.53–1.64) | 0.25 (0.15–0.40) | 0.35 (0.22–0.54) |
| Trauma and Orthopaedic Surgery | ST3+ | 0.30 (0.24–0.38) | 0.17 (0.11–0.25) | 0.59 (0.31–1.11) | 0.21 (0.14–0.32) | 0.16 (0.11–0.23) |

**Supplementary Table S4.** Offer odds ratios (95% CI) by country of primary medical qualification, compared with UK graduates. Programmes with at least 500 cumulative offers, 2021–2025.

| Programme | Level | EU/EEA vs UK | Rest of world vs UK | All IMG vs UK |
|---|---|---|---|---|
| Anaesthetics | ST1/CT1 | 0.16 (0.12–0.20) | 0.018 (0.015–0.022) | 0.030 (0.026–0.035) |
| Clinical Radiology | ST1/CT1 | 0.21 (0.17–0.27) | 0.086 (0.076–0.097) | 0.099 (0.088–0.111) |
| Core Psychiatry | ST1/CT1 | 0.22 (0.19–0.26) | 0.098 (0.092–0.105) | 0.10 (0.10–0.11) |
| Core Surgical Training | ST1/CT1 | 0.27 (0.23–0.31) | 0.087 (0.077–0.097) | 0.11 (0.10–0.13) |
| Emergency Medicine | ST1/CT1 | 0.18 (0.15–0.23) | 0.046 (0.041–0.052) | 0.055 (0.049–0.061) |
| General Practice | ST1/CT1 | 0.17 (0.16–0.19) | 0.12 (0.11–0.12) | 0.12 (0.12–0.13) |
| Histopathology | ST1/CT1 | 0.58 (0.41–0.83) | 0.22 (0.18–0.27) | 0.25 (0.21–0.30) |
| Internal Medicine | ST1/CT1 | 0.43 (0.38–0.47) | 0.25 (0.23–0.26) | 0.26 (0.25–0.28) |
| Obstetrics and Gynaecology | ST1/CT1 | 0.19 (0.16–0.24) | 0.052 (0.046–0.058) | 0.060 (0.054–0.067) |
| Ophthalmology | ST1/CT1 | 0.26 (0.17–0.38) | 0.043 (0.031–0.059) | 0.065 (0.050–0.083) |
| Paediatrics | ST1/CT1 | 0.35 (0.28–0.43) | 0.14 (0.12–0.15) | 0.15 (0.14–0.17) |
| Acute Internal Medicine | ST3+ | 0.59 (0.31–1.14) | 0.39 (0.32–0.48) | 0.40 (0.33–0.49) |
| Anaesthetics | ST3+ | 0.72 (0.53–0.99) | 0.53 (0.46–0.62) | 0.56 (0.49–0.65) |
| Cardiology | ST3+ | 0.49 (0.33–0.72) | 0.24 (0.20–0.29) | 0.26 (0.21–0.31) |
| Clinical Oncology | ST3+ | 0.45 (0.26–0.79) | 0.28 (0.23–0.36) | 0.29 (0.24–0.37) |
| Combined Infection Training | ST3+ | 0.42 (0.25–0.69) | 0.19 (0.15–0.24) | 0.21 (0.17–0.26) |
| Emergency Medicine | ST3+ | 0.54 (0.30–0.98) | 0.114 (0.090–0.145) | 0.12 (0.10–0.16) |
| Endocrinology and Diabetes Mellitus | ST3+ | 0.95 (0.54–1.67) | 0.38 (0.29–0.48) | 0.39 (0.31–0.50) |
| Gastroenterology | ST3+ | 0.41 (0.25–0.67) | 0.24 (0.20–0.30) | 0.25 (0.20–0.31) |
| General Surgery | ST3+ | 0.46 (0.31–0.66) | 0.18 (0.15–0.22) | 0.20 (0.17–0.24) |
| Geriatric Medicine | ST3+ | 0.39 (0.22–0.68) | 0.21 (0.17–0.27) | 0.22 (0.18–0.27) |
| Haematology | ST3+ | 0.35 (0.20–0.60) | 0.23 (0.18–0.28) | 0.23 (0.19–0.29) |
| Intensive Care Medicine | ST3+ | 0.44 (0.28–0.67) | 0.27 (0.22–0.32) | 0.28 (0.24–0.33) |
| Medical Oncology | ST3+ | 0.42 (0.25–0.70) | 0.28 (0.22–0.35) | 0.29 (0.23–0.36) |
| Paediatrics | ST3+ | 0.72 (0.34–1.56) | 0.30 (0.21–0.44) | 0.31 (0.21–0.45) |
| Psychiatry Subspecialities | ST3+ | 0.66 (0.48–0.90) | 0.43 (0.37–0.50) | 0.45 (0.39–0.52) |
| Respiratory Medicine | ST3+ | 0.34 (0.19–0.62) | 0.27 (0.23–0.33) | 0.28 (0.23–0.33) |
| Trauma and Orthopaedic Surgery | ST3+ | 0.46 (0.32–0.68) | 0.094 (0.076–0.116) | 0.12 (0.10–0.14) |

**Supplementary Table S5.** Offer odds ratios within broad ethnic groups, pooled over programmes with at least 500 cumulative offers. Crude: offers and applications summed across programmes (primary analysis). MH: Mantel–Haenszel odds ratio stratified by programme.

| Level | Comparison | Programmes | Crude OR (95% CI) | *p* | MH OR (95% CI) |
|---|---|---|---|---|---|
| ST1/CT1 | Bangladeshi vs Indian | 9 | 0.80 (0.74–0.86) | <0.001 | 0.73 (0.67–0.78) |
| ST1/CT1 | Pakistani vs Indian | 9 | 0.68 (0.65–0.71) | <0.001 | 0.63 (0.60–0.66) |
| ST1/CT1 | Other Asian vs Indian | 9 | 1.10 (1.05–1.16) | <0.001 | 1.05 (1.00–1.10) |
| ST1/CT1 | Caribbean vs African | 9 | 2.50 (2.15–2.90) | <0.001 | 2.62 (2.24–3.08) |
| ST1/CT1 | Other Black vs African | 9 | 1.75 (1.51–2.04) | <0.001 | 1.56 (1.34–1.83) |
| ST1/CT1 | Irish vs British | 9 | 0.75 (0.68–0.82) | <0.001 | 0.68 (0.62–0.75) |
| ST1/CT1 | Other White vs British | 9 | 0.27 (0.26–0.29) | <0.001 | 0.24 (0.22–0.25) |
| ST3+ | Bangladeshi vs Indian | 12 | 0.64 (0.52–0.78) | <0.001 | 0.73 (0.59–0.90) |
| ST3+ | Pakistani vs Indian | 12 | 0.57 (0.52–0.64) | <0.001 | 0.65 (0.58–0.73) |
| ST3+ | Other Asian vs Indian | 12 | 0.94 (0.83–1.06) | 0.297 | 0.96 (0.85–1.10) |
| ST3+ | Caribbean vs African | 11 | 1.92 (1.28–2.88) | 0.002 | 2.26 (1.50–3.39) |
| ST3+ | Other Black vs African | 12 | 0.77 (0.54–1.10) | 0.152 | 0.90 (0.61–1.33) |
| ST3+ | Irish vs British | 12 | 0.89 (0.73–1.10) | 0.284 | 0.90 (0.74–1.11) |
| ST3+ | Other White vs British | 12 | 0.56 (0.49–0.62) | <0.001 | 0.54 (0.47–0.61) |

**Supplementary Table S6.** Within-group offer odds ratios for the three largest entry-level programmes. Target and reference are the first- and second-named groups in each comparison.

| Programme | Comparison | Target offers/apps | Reference offers/apps | OR (95% CI) | *p* |
|---|---|---|---|---|---|
| Core Psychiatry | Bangladeshi vs Indian | 81/885 | 552/4,326 | 0.69 (0.54–0.88) | 0.003 |
| Core Psychiatry | Caribbean vs African | 14/112 | 642/8,995 | 1.93 (1.11–3.38) | 0.020 |
| Core Psychiatry | Irish vs British | 57/111 | 1,207/1,866 | 0.58 (0.39–0.85) | 0.005 |
| Core Psychiatry | Other Asian vs Indian | 254/1,829 | 552/4,326 | 1.10 (0.94–1.29) | 0.231 |
| Core Psychiatry | Other Black vs African | 16/159 | 642/8,995 | 1.41 (0.83–2.39) | 0.208 |
| Core Psychiatry | Other White vs British | 326/1,447 | 1,207/1,866 | 0.16 (0.14–0.19) | <0.001 |
| Core Psychiatry | Pakistani vs Indian | 438/5,116 | 552/4,326 | 0.64 (0.56–0.73) | <0.001 |
| General Practice | Bangladeshi vs Indian | 606/2,416 | 3,285/10,402 | 0.73 (0.66–0.80) | <0.001 |
| General Practice | Caribbean vs African | 156/336 | 4,422/17,756 | 2.61 (2.10–3.25) | <0.001 |
| General Practice | Irish vs British | 485/675 | 6,501/8,008 | 0.59 (0.50–0.71) | <0.001 |
| General Practice | Other Asian vs Indian | 2,032/6,017 | 3,285/10,402 | 1.10 (1.03–1.18) | 0.004 |
| General Practice | Other Black vs African | 162/471 | 4,422/17,756 | 1.58 (1.30–1.92) | <0.001 |
| General Practice | Other White vs British | 1,302/3,299 | 6,501/8,008 | 0.15 (0.14–0.17) | <0.001 |
| General Practice | Pakistani vs Indian | 3,230/13,491 | 3,285/10,402 | 0.68 (0.64–0.72) | <0.001 |
| Internal Medicine | Bangladeshi vs Indian | 150/542 | 1,352/3,912 | 0.72 (0.59–0.88) | 0.001 |
| Internal Medicine | Caribbean vs African | 28/83 | 428/1,923 | 1.78 (1.12–2.84) | 0.016 |
| Internal Medicine | Irish vs British | 167/317 | 2,596/4,546 | 0.84 (0.67–1.05) | 0.125 |
| Internal Medicine | Other Asian vs Indian | 567/1,748 | 1,352/3,912 | 0.91 (0.81–1.02) | 0.118 |
| Internal Medicine | Other Black vs African | 16/74 | 428/1,923 | 0.97 (0.55–1.70) | 0.902 |
| Internal Medicine | Other White vs British | 529/1,324 | 2,596/4,546 | 0.50 (0.44–0.57) | <0.001 |
| Internal Medicine | Pakistani vs Indian | 769/3,065 | 1,352/3,912 | 0.63 (0.57–0.70) | <0.001 |

**Supplementary Table S7.** Programme-level data for the ecological analysis: percentage of applications from graduates outside the UK and EU (ROW, 2021–2025) and offer odds ratios for Asian and Black applicants compared with White applicants.

| Programme | Level | ROW (%) | OR, Asian | OR, Black |
|---|---|---|---|---|
| Core Surgical Training | ST1/CT1 | 41.7 | 0.38 | 0.17 |
| Anaesthetics | ST1/CT1 | 46.0 | 0.12 | 0.027 |
| Internal Medicine | ST1/CT1 | 50.0 | 0.39 | 0.26 |
| Paediatrics | ST1/CT1 | 50.9 | 0.27 | 0.17 |
| Clinical Radiology | ST1/CT1 | 56.4 | 0.30 | 0.13 |
| Emergency Medicine | ST1/CT1 | 58.7 | 0.086 | 0.053 |
| General Practice | ST1/CT1 | 67.5 | 0.18 | 0.15 |
| Obstetrics and Gynaecology | ST1/CT1 | 69.5 | 0.15 | 0.059 |
| Core Psychiatry | ST1/CT1 | 78.2 | 0.14 | 0.090 |
| Anaesthetics | ST3+ | 15.8 | 0.70 | 0.58 |
| Intensive Care Medicine | ST3+ | 32.7 | 0.38 | 0.29 |
| Psychiatry Subspecialities | ST3+ | 47.9 | 0.58 | 0.41 |
| Trauma and Orthopaedic Surgery | ST3+ | 48.4 | 0.30 | 0.17 |
| Clinical Oncology | ST3+ | 50.5 | 0.52 | 0.48 |
| Geriatric Medicine | ST3+ | 52.4 | 0.26 | 0.23 |
| Medical Oncology | ST3+ | 53.5 | 0.55 | 0.42 |
| Respiratory Medicine | ST3+ | 58.4 | 0.31 | 0.28 |
| Cardiology | ST3+ | 62.4 | 0.38 | 0.32 |
| Haematology | ST3+ | 63.7 | 0.41 | 0.20 |
| General Surgery | ST3+ | 64.3 | 0.44 | 0.23 |
| Acute Internal Medicine | ST3+ | 73.8 | 0.37 | 0.35 |

**Supplementary Table S8.** Offer odds ratios with suppressed cells (fewer than five) set to 0, 2.5 (primary) and 5. Comparisons that fall below the 500-offer threshold under one of the bounds are omitted.

| Comparison | Programme | Level | OR (0) | OR (2.5) | OR (5) | Max change (%) | Significance changes |
|---|---|---|---|---|---|---|---|
| Asian vs White | Emergency Medicine | ST1/CT1 | 0.084 | 0.086 | 0.087 | 1.8 | No |
| Asian vs White | Anaesthetics | ST1/CT1 | 0.12 | 0.12 | 0.13 | 1.4 | No |
| Asian vs White | Core Psychiatry | ST1/CT1 | 0.14 | 0.14 | 0.14 | 0.0 | No |
| Asian vs White | Obstetrics and Gynaecology | ST1/CT1 | 0.15 | 0.15 | 0.15 | 2.3 | No |
| Asian vs White | General Practice | ST1/CT1 | 0.18 | 0.18 | 0.18 | 0.0 | No |
| Asian vs White | Paediatrics | ST1/CT1 | 0.26 | 0.27 | 0.27 | 1.3 | No |
| Asian vs White | Clinical Radiology | ST1/CT1 | 0.30 | 0.30 | 0.30 | 0.6 | No |
| Asian vs White | Core Surgical Training | ST1/CT1 | 0.38 | 0.38 | 0.38 | 0.0 | No |
| Asian vs White | Internal Medicine | ST1/CT1 | 0.39 | 0.39 | 0.39 | 0.2 | No |
| Asian vs White | Geriatric Medicine | ST3+ | 0.29 | 0.26 | 0.23 | 13.1 | No |
| Asian vs White | Trauma and Orthopaedic Surgery | ST3+ | 0.29 | 0.30 | 0.31 | 3.3 | No |
| Asian vs White | Respiratory Medicine | ST3+ | 0.32 | 0.31 | 0.29 | 5.6 | No |
| Asian vs White | Intensive Care Medicine | ST3+ | 0.35 | 0.38 | 0.41 | 7.3 | No |
| Asian vs White | General Surgery | ST3+ | 0.43 | 0.44 | 0.45 | 2.4 | No |
| Asian vs White | Clinical Oncology | ST3+ | 0.52 | 0.52 | 0.51 | 1.3 | No |
| Asian vs White | Psychiatry Subspecialities | ST3+ | 0.56 | 0.58 | 0.59 | 2.4 | No |
| Asian vs White | Anaesthetics | ST3+ | 0.68 | 0.70 | 0.71 | 2.6 | No |
| Black vs White | Anaesthetics | ST1/CT1 | 0.024 | 0.027 | 0.029 | 11.0 | No |
| Black vs White | Emergency Medicine | ST1/CT1 | 0.045 | 0.053 | 0.061 | 17.9 | No |
| Black vs White | Obstetrics and Gynaecology | ST1/CT1 | 0.054 | 0.059 | 0.064 | 9.5 | No |
| Black vs White | Core Psychiatry | ST1/CT1 | 0.088 | 0.090 | 0.092 | 2.5 | No |
| Black vs White | Clinical Radiology | ST1/CT1 | 0.12 | 0.13 | 0.14 | 12.8 | No |
| Black vs White | General Practice | ST1/CT1 | 0.15 | 0.15 | 0.15 | 0.0 | No |
| Black vs White | Core Surgical Training | ST1/CT1 | 0.15 | 0.17 | 0.19 | 13.6 | No |
| Black vs White | Paediatrics | ST1/CT1 | 0.16 | 0.17 | 0.19 | 10.9 | No |
| Black vs White | Internal Medicine | ST1/CT1 | 0.25 | 0.26 | 0.27 | 3.7 | No |
| Black vs White | Trauma and Orthopaedic Surgery | ST3+ | 0.14 | 0.17 | 0.19 | 20.2 | No |
| Black vs White | Geriatric Medicine | ST3+ | 0.25 | 0.23 | 0.21 | 11.0 | No |
| Black vs White | General Surgery | ST3+ | 0.21 | 0.23 | 0.25 | 11.4 | No |
| Black vs White | Respiratory Medicine | ST3+ | 0.26 | 0.28 | 0.30 | 7.8 | No |
| Black vs White | Intensive Care Medicine | ST3+ | 0.28 | 0.29 | 0.29 | 1.7 | No |
| Black vs White | Psychiatry Subspecialities | ST3+ | 0.39 | 0.41 | 0.42 | 4.0 | No |
| Black vs White | Clinical Oncology | ST3+ | 0.42 | 0.48 | 0.52 | 13.5 | No |
| Black vs White | Anaesthetics | ST3+ | 0.35 | 0.58 | 0.84 | 65.8 | Yes |
| Chinese vs White | Emergency Medicine | ST1/CT1 | 0.37 | 0.37 | 0.37 | 0.0 | No |

| Comparison | Programme | Level | OR (0) | OR (2.5) | OR (5) | Max change (%) | Significance changes |
|---|---|---|---|---|---|---|---|
| Chinese vs White | Anaesthetics | ST1/CT1 | 0.39 | 0.39 | 0.39 | 0.0 | No |
| Chinese vs White | Obstetrics and Gynaecology | ST1/CT1 | 0.60 | 0.60 | 0.60 | 0.0 | No |
| Chinese vs White | Paediatrics | ST1/CT1 | 0.63 | 0.63 | 0.63 | 0.0 | No |
| Chinese vs White | Core Surgical Training | ST1/CT1 | 0.83 | 0.83 | 0.83 | 0.0 | No |
| Chinese vs White | Clinical Radiology | ST1/CT1 | 0.83 | 0.83 | 0.82 | 0.6 | No |
| Chinese vs White | Core Psychiatry | ST1/CT1 | 0.97 | 0.97 | 0.97 | 0.0 | No |
| Chinese vs White | Internal Medicine | ST1/CT1 | 0.98 | 0.97 | 0.97 | 0.5 | No |
| Chinese vs White | General Practice | ST1/CT1 | 1.02 | 1.02 | 1.02 | 0.0 | No |
| Chinese vs White | Intensive Care Medicine | ST3+ | 0.46 | 0.54 | 0.63 | 17.8 | Yes |
| Chinese vs White | Geriatric Medicine | ST3+ | 0.61 | 0.54 | 0.48 | 13.1 | Yes |
| Chinese vs White | Trauma and Orthopaedic Surgery | ST3+ | 0.35 | 0.59 | 0.91 | 65.4 | Yes |
| Chinese vs White | Clinical Oncology | ST3+ | 0.46 | 0.63 | 0.90 | 41.7 | Yes |
| Chinese vs White | Psychiatry Subspecialities | ST3+ | 0.71 | 0.71 | 0.71 | 0.0 | No |
| Chinese vs White | Respiratory Medicine | ST3+ | 1.00 | 0.93 | 0.87 | 7.3 | No |
| Chinese vs White | Anaesthetics | ST3+ | 0.97 | 0.97 | 0.97 | 0.2 | No |
| Chinese vs White | General Surgery | ST3+ | 1.04 | 1.03 | 1.01 | 1.3 | No |
| Mixed vs White | Obstetrics and Gynaecology | ST1/CT1 | 0.15 | 0.19 | 0.22 | 21.3 | No |
| Mixed vs White | Emergency Medicine | ST1/CT1 | 0.15 | 0.19 | 0.22 | 22.5 | No |
| Mixed vs White | Core Psychiatry | ST1/CT1 | 0.20 | 0.21 | 0.21 | 2.1 | No |
| Mixed vs White | Anaesthetics | ST1/CT1 | 0.18 | 0.21 | 0.24 | 15.6 | No |
| Mixed vs White | General Practice | ST1/CT1 | 0.23 | 0.23 | 0.23 | 0.0 | No |
| Mixed vs White | Clinical Radiology | ST1/CT1 | 0.16 | 0.26 | 0.35 | 56.4 | No |
| Mixed vs White | Paediatrics | ST1/CT1 | 0.25 | 0.28 | 0.32 | 13.5 | No |
| Mixed vs White | Core Surgical Training | ST1/CT1 | 0.29 | 0.33 | 0.38 | 15.1 | No |
| Mixed vs White | Internal Medicine | ST1/CT1 | 0.47 | 0.47 | 0.47 | 0.1 | No |
| Mixed vs White | Trauma and Orthopaedic Surgery | ST3+ | 0.003 | 0.21 | 0.46 | 6383.5 | No |
| Mixed vs White | Geriatric Medicine | ST3+ | 0.031 | 0.22 | 0.52 | 598.7 | No |
| Mixed vs White | Respiratory Medicine | ST3+ | 0.053 | 0.25 | 0.48 | 367.5 | No |
| Mixed vs White | General Surgery | ST3+ | 0.10 | 0.26 | 0.45 | 169.1 | No |
| Mixed vs White | Clinical Oncology | ST3+ | 0.047 | 0.34 | 0.83 | 627.8 | Yes |
| Mixed vs White | Intensive Care Medicine | ST3+ | 0.13 | 0.36 | 0.73 | 188.3 | Yes |
| Mixed vs White | Psychiatry Subspecialities | ST3+ | 0.59 | 0.67 | 0.75 | 12.1 | Yes |
| Mixed vs White | Anaesthetics | ST3+ | 0.59 | 0.80 | 1.06 | 35.3 | Yes |
| Other vs White | Anaesthetics | ST1/CT1 | 0.062 | 0.062 | 0.062 | 0.0 | No |
| Other vs White | Emergency Medicine | ST1/CT1 | 0.082 | 0.082 | 0.082 | 0.0 | No |
| Other vs White | Obstetrics and Gynaecology | ST1/CT1 | 0.12 | 0.12 | 0.12 | 0.0 | No |
| Other vs White | Paediatrics | ST1/CT1 | 0.21 | 0.21 | 0.21 | 0.0 | No |

| Comparison | Programme | Level | OR (0) | OR (2.5) | OR (5) | Max change (%) | Significance changes |
|---|---|---|---|---|---|---|---|
| Other vs White | Clinical Radiology | ST1/CT1 | 0.22 | 0.22 | 0.22 | 0.6 | No |
| Other vs White | Core Psychiatry | ST1/CT1 | 0.24 | 0.24 | 0.24 | 0.0 | No |
| Other vs White | General Practice | ST1/CT1 | 0.27 | 0.27 | 0.27 | 0.0 | No |
| Other vs White | Core Surgical Training | ST1/CT1 | 0.29 | 0.29 | 0.29 | 0.0 | No |
| Other vs White | Internal Medicine | ST1/CT1 | 0.33 | 0.33 | 0.33 | 0.0 | No |
| Other vs White | Trauma and Orthopaedic Surgery | ST3+ | 0.17 | 0.16 | 0.15 | 4.2 | No |
| Other vs White | Geriatric Medicine | ST3+ | 0.17 | 0.19 | 0.21 | 11.9 | No |
| Other vs White | Intensive Care Medicine | ST3+ | 0.32 | 0.32 | 0.32 | 0.0 | No |
| Other vs White | General Surgery | ST3+ | 0.33 | 0.32 | 0.32 | 1.3 | No |
| Other vs White | Respiratory Medicine | ST3+ | 0.37 | 0.35 | 0.32 | 7.3 | No |
| Other vs White | Clinical Oncology | ST3+ | 0.50 | 0.47 | 0.45 | 5.8 | No |
| Other vs White | Anaesthetics | ST3+ | 0.50 | 0.50 | 0.50 | 0.2 | No |
| Other vs White | Psychiatry Subspecialities | ST3+ | 0.66 | 0.66 | 0.66 | 0.0 | No |

Preprint, revised 10th November 2026
